# CovRNN—A recurrent neural network model for predicting outcomes of COVID-19 patients: model development and validation using EHR data

**DOI:** 10.1101/2021.09.27.21264121

**Authors:** Laila Rasmy, Masayuki Nigo, Bijun Sai Kannadath, Ziqian Xie, Bingyu Mao, Khush Patel, Yujia Zhou, Wanheng Zhang, Angela Ross, Hua Xu, Degui Zhi

## Abstract

**Background:** Predicting outcomes of COVID-19 patients at an early stage is critical for optimized clinical care and resource management, especially during a pandemic. Although multiple machine learning models have been proposed to address this issue, based on the need for extensive data pre-processing and feature engineering, these models have not been validated or implemented outside of the original study site.

**Methods:** In this study, we propose CovRNN, recurrent neural network (RNN)-based models to predict COVID-19 patients’ outcomes, using their available electronic health record (EHR) data on admission, without the need for specific feature selection or missing data imputation. CovRNN is designed to predict three outcomes: in-hospital mortality, need for mechanical ventilation, and long length of stay (LOS >7 days). Predictions are made for time-to-event risk scores (survival prediction) and all-time risk scores (binary prediction). Our models were trained and validated using heterogeneous and de-identified data of 247,960 COVID-19 patients from 87 healthcare systems, derived from the Cerner® Real-World Dataset (CRWD). External validation was performed using three test sets (approximately 53,000 patients). Further, the transferability of CovRNN was validated using 36,140 de-identified patients’ data derived from the Optum® de-identified COVID-19 Electronic Health Record v. 1015 dataset (2007–2020).

**Findings:** CovRNN shows higher performance than do traditional models. It achieved an area under the receiving operating characteristic (AUROC) of 93% for mortality and mechanical ventilation predictions on the CRWD test set (vs. 91·5% and 90% for light gradient boost machine (LGBM) and logistic regression (LR), respectively) and 86.5% for prediction of LOS > 7 days (vs. 81·7% and 80% for LGBM and LR, respectively). For survival prediction, CovRNN achieved a C-index of 86% for mortality and 92·6% for mechanical ventilation. External validation confirmed AUROCs in similar ranges.

**Interpretation:** Trained on a large heterogeneous real-world dataset, our CovRNN model showed high prediction accuracy, good calibration, and transferability through consistently good performance on multiple external datasets. Our results demonstrate the feasibility of a COVID-19 predictive model that delivers high accuracy without the need for complex feature engineering.

## INTRODUCTION

Coronavirus disease 2019 (COVID-19) is an infectious disease caused by the severe acute respiratory syndrome coronavirus 2 (SARS-CoV-2), which emerged in December 2019.^1^ By the end of August 2021, there were more than 215 million confirmed COVID-19 infections worldwide and more than 630,000 deaths in the United States alone.^2^ Further, there have been around three million hospital admissions recorded since August 2020.^2^ During the peaks of the pandemic waves, many states have reported near-capacity hospital and intensive care unit (ICU) utilization. Accurate prediction of the future clinical trajectories of COVID-19 patients at the time of admission is crucial for clinical decision making and enables efficient allocation of resources. Indeed, a number of models for the prediction of COVID-19 outcomes have been developed. Wynants et al.^3^ reviewed 107 prognostic models as of July 1, 2020. The most common issue highlighted in this study is the high risk of bias associated with the reviewed models, due to either the small, locally sourced training dataset and the high risk of model overfitting or the lack of model calibration or external validation.^4,5^ To provide an updated survey of the literature, we conducted a Scopus and PubMed search for COVID-19 outcomes prediction articles published between July 2020 and the end of April 2021, using the keywords “COVID electronic health record (‘mortality’ or ‘ventilator’ or ‘length of stay’ or ‘real-time’) prediction.” The literature search retrieved a total of 264 unique articles, and upon review, we found 31 studies that describe the development and validation of machine learning predictive models for COVID-19 patients’ prognosis after admission. Out of the 31 studies, only two studies^6,7^ involved training the proposed models on more than 20,000 patients. Both models are based on a small set of specific features and need a laborious data preprocessing and feature engineering process that limits the transferability, reliability, and sustainability of the models.

In this study, we aim to develop an accurate and transferrable model for COVID-19 patients’ outcomes on admission that include in-hospital mortality (iMort), need for mechanical ventilation during the stay (mVent), and hospital stay longer than one week (pLOS). Our model, CovRNN, utilizes a gated RNN architecture, proven to be effective in modeling patients’ electronic health records (EHR) data.^8–12^ To maximize transferability, CovRNN uses readily available structured EHR without any need for specific feature selection, feature engineering, or missing value imputation. For iMort and mVent prediction tasks, CovRNN predicts a time-to-event risk score that can be interpreted as a binary prediction with a time horizon (survival prediction) and an all-time risk score (binary prediction).

CovRNN was trained on a cloud-based, large heterogeneous dataset of 243,785 de-identified patients’ data derived from 85 health systems available through the Cerner® Real-World COVID-19 Q3 Dataset (CRWD), hosted on the Cerner HealtheDataLab™. We evaluated our models on four test sets extracted from the CRWD and the Optum® de-identified COVID-19 Electronic Health Record v.1015 dataset (2007–2020). Each test set is different in size and represents a different use case so that we can evaluate the cross-hospital generalizability and model transferability between different EHR data sources. We also reported the results of subgroup analysis and ablation studies for a better understanding of the model’s performance. In addition, we utilized the integrated gradient technique^13^ to expose the factors of the CovRNN predictions that can enhance the interpretability.

To the best of our knowledge, CovRNN is the first COVID-19 outcome prediction model that can simultaneously achieve the following: (i) accurately predict different COVID-19 patients’ outcomes on admission, and (ii) use readily available structured EHR in its raw categorical format without the need for specific feature selection or missing value imputation. In addition, the prospective compliance of CovRNN is evaluated against quality standards, including the transparent reporting of individual prognosis or diagnosis (TRIPOD) and the prediction model risk of bias assessment tool (PROBAST). We also showed the value added of the fine-tuning utility of CovRNN and how it can be used to improve the model’s prediction accuracy. Such utility can be further used for the continuous improvement of the model as per good machine learning practice (GMLP) recommendations to secure the model’s reliability and sustainability. The source code of our model is publicly available to enable its applications and further evaluation by other researchers.

## METHODS

### Datasets and Cohort Description

We extracted our main training cohort from the CRWD hosted on the Cerner HealtheDataLab™, a cloud-based de-identified patients’ dataset that included up to five years of historic clinical data for COVID-19 patients from 87 health systems, as of the end of September 2020. The CRWD included only patients who had a minimum of one emergency or inpatient encounter with a diagnosis code that could be associated with COVID-19 exposure or infection, or a positive result for a COVID-19 laboratory test. Further description of the CRWD is available in Supplementary Material A. In our study, we predefined our prediction point as the first COVID-19 hospitalization admission day to an emergency, observation, or in-patient unit, and we refer to this point as the *index date* (Figure 1). We thereby excluded all patients who had no recorded clinical information on or before the index date as well as patients who stayed for less than one day, as described in Supplementary Material B We also excluded patients who had inconsistent dates, such as discharge dates before the hospitalization start date, as well as patients who were readmitted later and presented different outcomes. Our cohort included 247,960 patients, from which we held out two hospitals’ data for external validation. The remaining 243,785 patients’ data were split into training, validation, and test sets, with the ratio of 7:1:2. All of our reported CRWD results were on the held-out test set of 48,781 patients from the 85 health systems. For external validation, we evaluated the model on two randomly selected held-out hospitals from the CRWD, Hospital 1 from the south region, with 3,469 patients, and Hospital 2 from the west region, with 706 patients.

**Figure 1.**
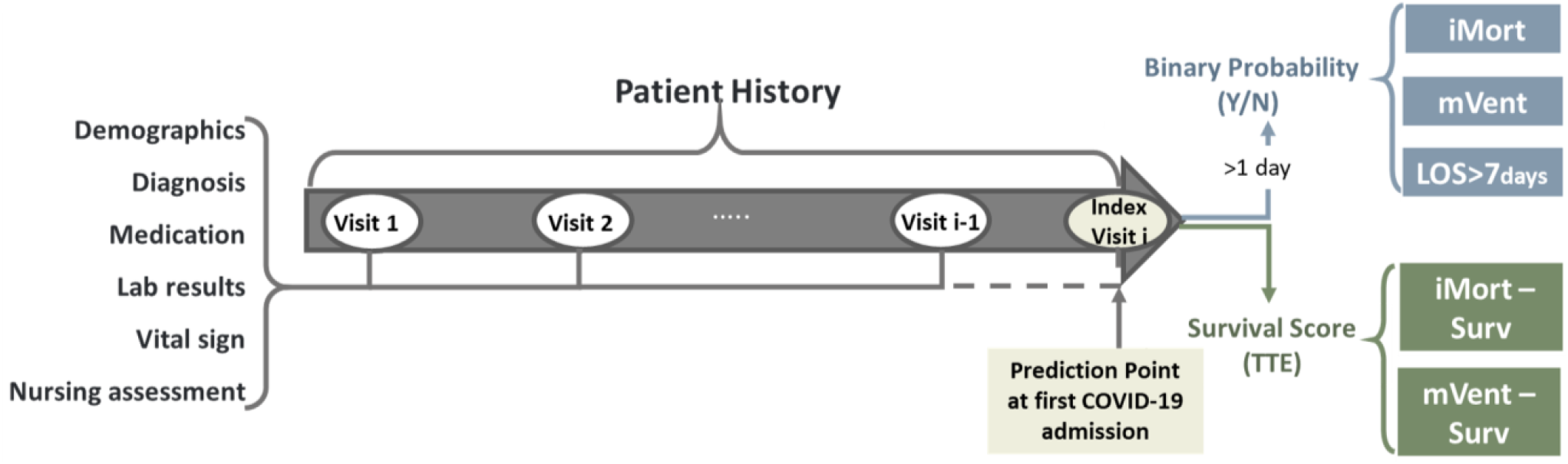
CovRNN prediction tasks.

For further external validation outside of the CRWD, we extracted a cohort of 36,140 de-identified patients’ data derived from the Optum® de-identified COVID-19 Electronic Health Record dataset v.1015 dataset (2007-2020), which we refer to as “OPTUM” cohort. Further description of the Optum® dataset, along with differences and commonalities between the CRWD and OPTUM cohorts, are available in Supplementary Material A.

### Data Preparation

We kept our data curation to the minimum level, as we describe below, to facilitate the transferability of our trained models among different datasets. We extracted all patient information on or before the date of their first hospital admission with COVID-19, including demographics, diagnosis, medication, procedures, laboratory results, and observations. To facilitate interoperability, we utilized standard terminologies in common use, such as ICD-9, ICD-10, and SNOMED-CT for diagnosis; LOINC and SNOMED-CT for laboratory results and observations; Multum drug identifiers and categories for medications; and CPT-4, HCPCS, ICD-9 PCs, and ICD-10 PCs for procedures. Such standard terminologies are readily available in the majority of EHR systems. In cases for which a coding system, such as Multum codes for medication, is not used, pre-existing mapping tools are available^14^ that can be used to convert NDC medication codes to corresponding Multum information.

The majority of our features, such as diagnosis, medications, and procedures, were categorical. We converted numeric variables, such as laboratory results, to categorical variables as follows. For the CRWD, we used the “below normal low, normal, or above normal high” interpretation value provided in the CRWD rather than the actual numerical value; for OPTUM, we defined the result categories based on the corresponding normal result ranges. By doing, so we can further convert our input to either multi-hot or embedding matrices to feed to our models. Based on our previous study,^15^ we decided to use the clinical information in the coding standards with which it was recorded, as the normalization of those codes to a more unified terminology provides minimal gain.^15^ Further details of our data curation are available in Supplementary Material B. Our data curation pipeline is available at https://github.com/ZhiGroup/CovRNN.

### Outcomes of Prediction Tasks

Our tasks include the prediction of COVID-19 patients’ in-hospital mortality (iMort), need for mechanical ventilation (mVent), and prolonged length of stay (pLOS), on admission. For iMort event definition, we relied on the pre-assigned mortality flags on CRWD along with the “expired” encounter discharge disposition to confirm in-hospital mortality and identify the date of death. The iMort event definition was slightly different on the OPTUM data because there was no clear discharge disposition that indicated patient in-hospital death. We instead used the date of death and compared it against the hospitalization discharge date to assign the proper label. For mVent, we used mainly relevant mechanical ventilation procedure codes to define the outcome. In addition, with the CRWD, we used other relevant observations and recorded ventilator settings, not only to identify the instance of the event but also to identify the earliest time of the event (Supplementary Table 1). For iMort and mVent prediction tasks, we trained survival and binary classification-based prediction models. For survival analysis, we defined the time to event as the number of days between our index date and the earliest date that indicated the occurrence of the event, either a laboratory result or a recorded procedure for mVent or the discharge date of iMort. We used the hospitalization discharge date as the censoring time. We defined pLOS as a binary indicator for hospitalizations longer than 7 days, as the median length of stay (LOS) in the CRWD and OPTUM cohorts were 3 and 5, respectively, and we trained only a binary classification model for the pLOS task.

We used the area under the receiver operating characteristic (AUROC) as the main model discriminative performance metrics for the binary prediction models. We also reported other clinically relevant metrics, including specificity at 95% sensitivity, the area under the precision-recall curve (AUPRC), and the sensitivity and specificity at optimum threshold, defined using the validation set. For survival analysis, we reported the concordance index^16^ (c-index) as our main evaluation metric. In addition, we used the predicted score and calculated the AUROC at different periods.

### Models

CovRNN models were based on a gated type of RNN, namely a gated recurrent unit (GRU), which is known for being an efficient sequential deep learning architecture for clinical event predictions.^10,17^ Our models were designed to consume all demographics, diagnoses, medications, procedures, laboratory results, and other clinical event information readily available in the EHR before or on the index date to predict patient outcomes, without the need for specific feature selection or missing value imputation, for convenience and practicality. CovRNN also consumes the time difference between visits for a better temporal representation of patient history, which is known to slightly improve the prediction accuracy.^18,19^ For binary classification tasks, we compared CovRNN against traditional machine learning algorithms, such as logistic regression (LR)^20^ and light gradient boost machine (LGBM).^21^ For survival prediction, we utilized the DeepSurv^22^ architecture, while replacing the multiple layer perceptron (MLP) with GRU for better sequential information modeling. We were unable to adequately compare against machine learning survival models, such as random survival forest (RSF), for computational resource restrictions on the Cerner HealtheDataLab™, especially with the increased number of covariates and large training set size. Any version of the RSF model runnable on Cerner HealtheDataLab™ had a very small number of iterations/trees that led to poor and unreliable results; therefore, we decided not to report these results. Further implementation details are available in Supplementary Material C.

### Experiments

For model development, we trained our models on 70% of the CRWD 85 hospitals’ data (training set) and used 10% (validation set) to determine the best model trained, while controlling for overfitting. We reported the model performance on the remaining 20% held out for external validation (multi-hospital test set).

For further external validation, we used two additional levels of test sets (Figure 2). First, two hospitals’ datasets were fully held out and used only to evaluate the cross-hospital generalizability: Hospital 1 (*n* = 3,469), from the south region, and Hospital 2 (*n* = 706), from the west region. Second, to evaluate the transferability of the CovRNN models across different EHR data sources, we used the OPTUM data set. Although the models can be directly used and evaluated on the OPTUM cohort, it is recommended to fine-tune the transferred model on a sample data of the destination, for two reasons: (a) Some clinical code distribution may vary or be newly presented at the destination data source; thus, during the model’s fine-tuning, these codes would get introduced to the model and become embedded closer to codes of similar meaning; and (b) The definition of the outcome variables can be slightly different, given the limitations of each data source; for example, the mVent outcome was defined mainly in the OPTUM cohort, using only the procedure codes, whereas the CRWD used procedure codes and additional clinical event results. Therefore, to evaluate the value added of the models’ fine-tuning, we transferred the best models trained on the CRWD and evaluated it on the OPTUM cohort before and after fine-tuning. We also compared the performance of the fine-tuned models against new models that were trained only on the same OPTUM data used for fine-tuning.

**Figure 2.**
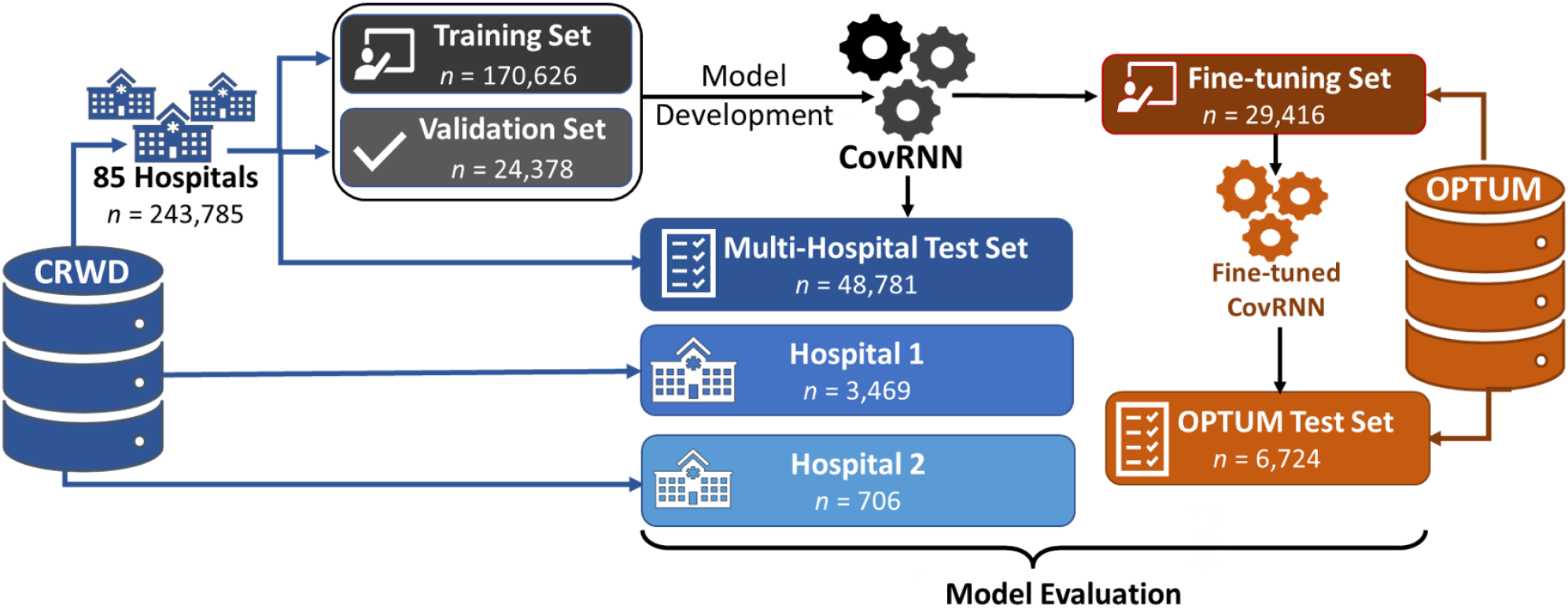
Model development and external validation datasets.

We designed our additional experiments using the CRWD multi-hospital test set as follows. The first experiment was an ablation experiment to evaluate the added value for each clinical data category, starting with diagnosis information, followed by medication, medication categories, laboratory tests, assessments results, procedures, and, lastly, demographics. The second experiment was a subgroup analysis to evaluate the validity of CovRNN for new patients who were admitted to the hospital for the first time and had no past medical history available in their records. Therefore, we compared the performance of CovRNN models on a modified version of the multi-hospital test set that includes only the last (index) visit information against the original full-history multi-hospital test set. For a better understanding of the impact of any possible label leakage during model training, we conducted our third experiment using the binary classification CovRNN for the mVent task. As our cohort definition excluded any patients with a stay of less than one day, our cohort did not include any patients who died within one day of admission. Nevertheless, nearly half of the intubated patients were intubated on their first day. Therefore, we evaluated the effect of excluding such patients, which we refer to as a “restricted” dataset, and compared the performance against our original “full” cohort. In addition to the above-mentioned three experiments, we conducted subgroup analysis for patients in regard to different age groups, races, baseline comorbidities, and geographical regions.

### Models Interpretation

For CovRNN prediction interpretation, we used the integrated gradient technique^13^ to expose the factors that contribute to the personalized model predictions. We used the integrated gradients technique due to its good theoretical properties, such as implementation invariance and completeness, and its implementation simplicity; as compared with methods such as layer-wise relevance propagation (LRP) or DeepLIFT, it does not require modification of the gradient backpropagation process and can viewed as a deterministic and computationally efficient approximation of the gradient Shapley Additive Explanations (SHAP). This is unlike LR- and LGBM-based models, in which the existing interpretation utilities provide fixed feature-level importance, by using either the LR coefficients or the LGBM feature importance scores. For RNN-based models, we can achieve a more personalized explanation that shows the contribution scores for each code at each patient visit. For the preliminary evaluation, we extracted 20 random sample patients and presented their predicted risk scores as well as the contribution score assigned for each medical event and asked infectious disease specialists to evaluate its relevance.

## RESULTS

The description of the overall CRWD and OPTUM cohorts, presented in Table 1, shows that the prevalence of the outcome variables varies across the different data sources. The OPTUM cohort shows about twice the prevalence of in-hospital mortality and evidence of mechanical ventilation. In addition, the median length of stay is longer in the OPTUM cohort. Supplementary Table 2 includes further details of each subset.

**Table 1.** Descriptive statistics for CRWD and OPTUM extracted cohorts.

| Characteristics | CRWD<br>*n* = 247,960 | OPTUM<br>*n* = 36,140 |
| --- | --- | --- |
| Age at the index visit |  |  |
| Median (IQR) | 57 (36–72) | 60 (44–72) |
| Gender |  |  |
| Female | 130,540 (52.6%) | 18,237 (50.5%) |
| Male | 116,653 (47.0%) | 17,885 (49.5%) |
| Race & Ethnicity |  |  |
| Caucasian | 168,606 (68.0%) | 19,704 (54.5%) |
| African American | 36,762 (14.8%) | 7,836 (21.7%) |
| Asian | 5,494 (2.2%) | 930 (2.6%) |
| American Indian / Alaska Native | 4,285 (1.7%) | NA |
| Hispanic | 72,068 (29.1%) | 5,782 (16.0%) |
| Comorbidities |  |  |
| Hypertension (HTN) | 114,387 (47.7%) | 22,035 (61.0%) |
| Diabetes (DM) | 64,023 (26.7%) | 12,942 (35.8%) |
| Congestive Heart Failure (CHF) | 36,040 (15.0%) | 6,568 (18.2%) |
| Chronic Kidney Disease (CKD) | 34,789 (14.5%) | 7,517 (20.8%) |
| Cancer | 19,145 (8.0%) | 5,094 (14.1%) |
| Outcomes |  |  |
| Mortality | 13,607 (5.5%) | 4,831 (13.4%) |
| Median TTE | 8 (4–16) | 5 (3–10) |
| Mechanical ventilation | 33,505 (13.5%) | 9,582 (26.5%) |
| Intubated on first day | 17,811 (7.2%) | 4,466 (12.4%) |
| Median TTE | 2 (1–5) | 3 (2–7) |
| Length of stay |  |  |
| Median (IQR) | 3 (1–6) | 5 (3–10) |
| Total number of unique features | 123,642 | 67,128 |
| Number of Healthcare Systems | 87 | 197 |

IQR: interquartile range, TTE: time to event in days

CovRNN binary classification models achieved an AUROC of 93% for iMort and mVent tasks on the CRWD test set (vs. 91·5% and 90% for LGBM and LR, respectively) and 86·5% for pLOS (vs. 81·7% and 80% for LGBM and LR, respectively) (Table 2). External validation on held-out hospitals data showed AUROC ranges of 91·5%–97% for iMort and mVent binary predictions. pLOS prediction task showed AUROC 87·2% and 88·3% for Hospital 1 and Hospital 2, respectively (Table 2). External validation on the OPTUM cohort showed an AUROC after fine-tuning of 91·3%, 91·5%, and 81·0% for iMort, mVent, and pLOS, respectively (Table 3). Additional metrics, including specificity at 95% sensitivity, AUPRC, and sensitivity and specificity at the optimum threshold, are presented in Supplementary Table 2.

**Table 2.** Model performance on different CRWD test sets.

|  | *n* | In-hospital Mortality (iMort) |  |  |  | Mechanical Ventilation (mVent) |  |  |  | Stay > 7 days (pLOS) |  |  |
| --- | --- | --- | --- | --- | --- | --- | --- | --- | --- | --- | --- | --- |
|  |  | LR | LGBM | Cov-RNN | CovRNN –SURV* | LR | LGBM | Cov-RNN | CovRNN –SURV* | LR | LGBM | Cov-RNN |
| **Multi-hospital Test Set** | 48,781 | 90.3 | 91.5 | 93.0 | 86.0 | 89.5 | 91.2 | 92.9 | 92.6 | 80.0 | 81.7 | 86.5 |
| **Hospital 1** | 3,469 | 88.8 | 91.0 | 91.8 | 86.0 | 86.7 | 88.4 | 91.5 | 90.8 | 77.3 | 78.5 | 87.2 |
| **Hospital 2** | 706 | 94.6 | 95.1 | 97.0 | 91.6 | 93.5 | 95.6 | 96.0 | 93.8 | 80.9 | 84.3 | 88.3 |

\* *CovRNN–SURV* shown values are the *c*-index, other models are showing AUROC for binary classifications evaluation

**Table 3.**
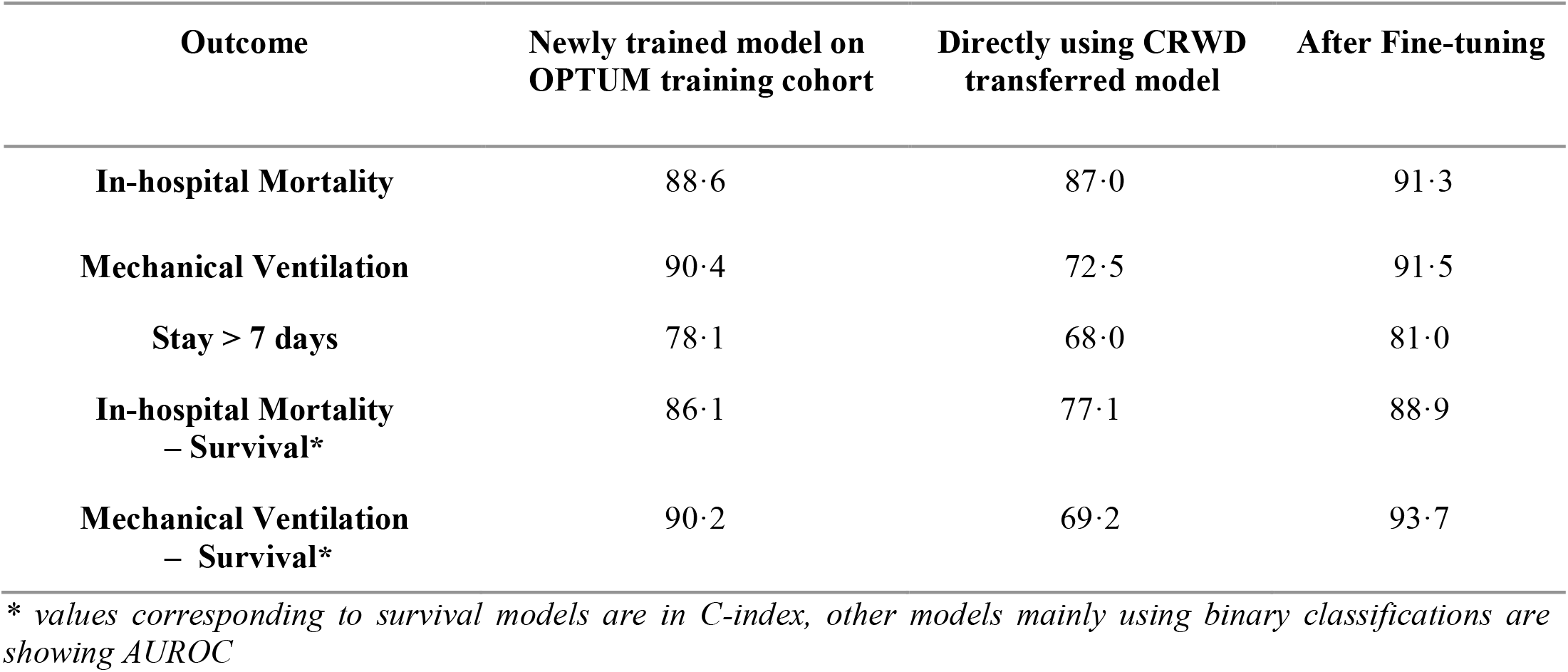
CovRNN models trained on CRWD performance on OPTUM before and after fine-tuning.

CovRNN survival models’ evaluation on the CRWD test set achieved a c-index of 86·0 for iMort and 92·6 for mVent (Table 2). Using the survival models to predict patient risk to develop the event at a certain time point within the period between Day 1 and Day 60 showed an AUROC range from 93·6% to 88·8% for iMort and from 95·5% to 91·4% for mVent. Similarly, the survival models showed a c-index range from 86·5 to 93·8 for iMort and mVent tasks on the held-out hospitals data (Table 2, Figure 3a) and the OPTUM test set after fine-tuning (Table 3, Figure 3b).

**Figure 3.**
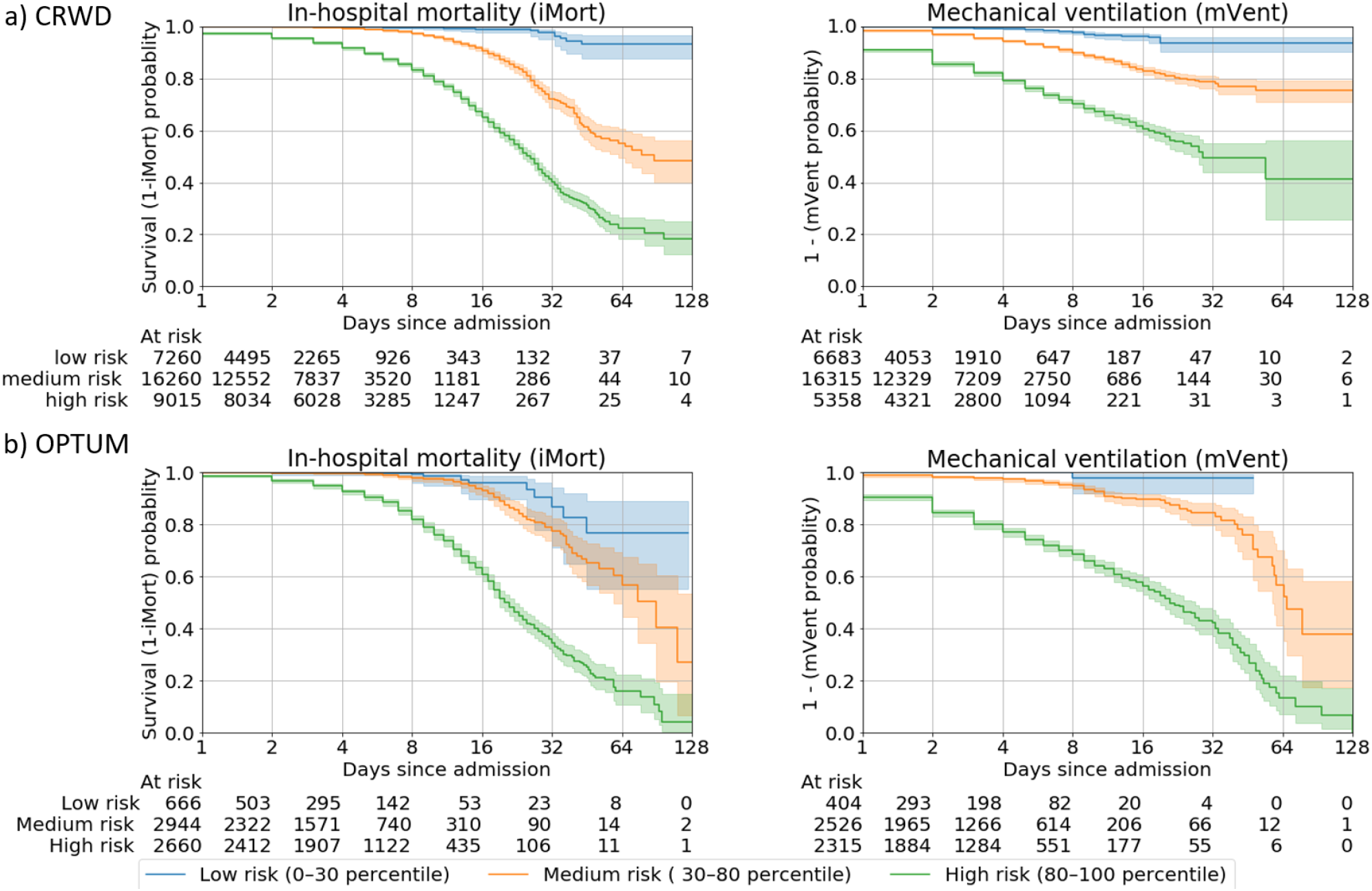
Kaplan-Meier (KM) curves that show the stratified survival analysis. Stratification of patients is according to their predicted survival score over time in days since admission. Shaded areas indicate 95% CIs calculated on the logarithmic scale from the standard errors of the Kaplan-Meier estimator with the center values as corresponding to the Kaplan–Meier estimate.

For CovRNN binary classification and survival models, the transferred models consistently achieved better performance after fine-tuning compared to training new models on the OPTUM data. Our transferred binary classification models after fine-tuning showed AUROCs of 91·3%, 91·2%, and 81% for iMort, mVent, and pLOS prediction tasks, respectively, compared to 88·7%, 90·4%, and 78·1% for newly trained models (Table 3). Similarly, transferred survival models showed a c-index range from 88·9% to 93·7% after fine-tuning for iMort and mVent versus 86·1% to 90·2% for newly trained models (Table 3).

Our first ablation experiment showed that each clinical data category contributes to an increase in the model prediction accuracy. For example, the addition of medication or laboratory results contributed to a 4% increase in the prediction accuracy for iMort or mVent tasks, not only for deep learning-based models but also for LR and LGBM (Table 4). Our second experiment results showed that the use of the full patient history continuously had a better performance than using only the last (index) visit information only (Table 5). Notably, the models’ performance remains acceptable without the use of previous medical records, especially for the iMort and mVent tasks, which show an AUROC of 92%. (Table 5). For the pLOS task, there is a higher decrease in the prediction accuracy, by 3.5%. Interestingly, this decrease in pLOS prediction accuracy also aligns with the higher prediction accuracy improvement of 6% for CovRNN models compared to LR-based model for the pLOS task versus an improvement of only 3% for the iMort and mVent tasks. In our third experiment, we found that training a version of the mVent binary prediction model, using the “restricted” training set, reduced the prediction accuracy by 3% on the full test set for CovRNN and 5% for LR and LGBM (Table 6). CovRNN performance remains constant on the “restricted” test set, regardless of which cohort it was originally trained on.

**Table 4.** Experiment 1- Ablation study investigating the impact of different data categories.

| Characteristics | Number of<br>covariates | Mortality (iMort) |  |  | Ventilator Use (mVent) |  |  |
| --- | --- | --- | --- | --- | --- | --- | --- |
|  |  | LR | LGBM | CovRNN | LR | LGBM | CovRNN |
| **Diagnosis only**<br>(ICD-9 / ICD-10 / SNOMED CT) | 49,074 | 77·6 | 83·8 | 85·9 | 75·5 | 80·9 | 83·7 |
| **Diagnosis + Medication**<br>(*Multum dNUM & Multilevel categories*) | 52,177 | 81·4 | 86·6 | 88·3 | 80·8 | 85·2 | 87·8 |
| **Diagnosis + Medication + Lab results**<br>(*LOINC codes with categorical results/*<br>*interpretations*) | 80,203 | 85·3 | 90·1 | 92·1 | 84·6 | 89·2 | 91·4 |
| **Diagnosis + Medication + Lab and**<br>**other assessments results + Procedures**<br>(*CPT-4, HCPCS, SNOMED CT,*<br>*ICD-9/10Pcs*) | 125,821 | 85·4 | 90·7 | 92·5 | 86·2 | 90·1 | 92·1 |
| **Diagnosis + Medication + Lab and other**<br>**assessments results + Procedures +**<br>**Demographics**<br>(*Race, gender, age, location*) | 125,917 | 86·4 | 90·9 | 92·7 | 86·2 | 90·1 | 92·3 |

LR: logistic regression, LGBM: light gradient boost machine

**Table 5.**
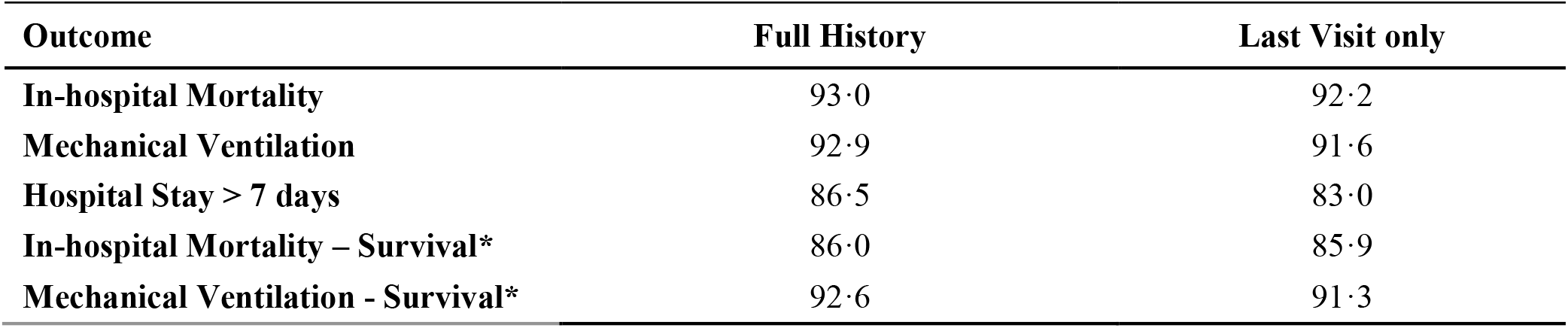
Experiment 2 - Model performance using only last visit data versus using full patient history.

**Table 6.** Experiment 3 - Effect of label leakage on the need for mechanical ventilation prediction.

| Trained | Full Test Data |  |  | Restricted Test Data |  |  |
| --- | --- | --- | --- | --- | --- | --- |
|  | LR | LGBM | CovRNN | LR | LGBM | CovRNN |
| **Full Data** | 89·5 | 91·2 | 92·9 | 81·5 | 82·8 | 85·9 |
| **Restricted Data** | 83·9 | 86·6 | 90·0 | 81·8 | 83·8 | 86·0 |

The subgroup analysis showed that prediction accuracy remains mainly consistent among different comorbidities, age groups, races, and regions. The most notable trend is that the prediction accuracy is better among the younger population (Figure 4). In addition, CovRNN binary classification models showed good calibration without sacrificing high prediction accuracy, as shown in the calibration plots (Supplementary Figure 4). In Supplementary Figure 5, we present a sample visualization that shows the integrated gradient-based explanation of the CovRNN model’s true positive prediction for a pLOS case. The information in the figure, however, is based on a sample and not the full patient data; the full data for a subset of more than 20 patients for each prediction task were presented to an infectious disease specialist, who found it informative. Further evaluation of the model explanation is warranted, taking into consideration that the evaluation of such personalized explanations, whereby the same clinical code can have different contribution scores at each patient and visit level, given the different context, is laborious. To demonstrate our efforts to abide by transparent reporting standards, we provide the TRIPOD and PROBAST assessments as Supplementary Material D and Supplementary Material E, respectively.

**Figure 4.**
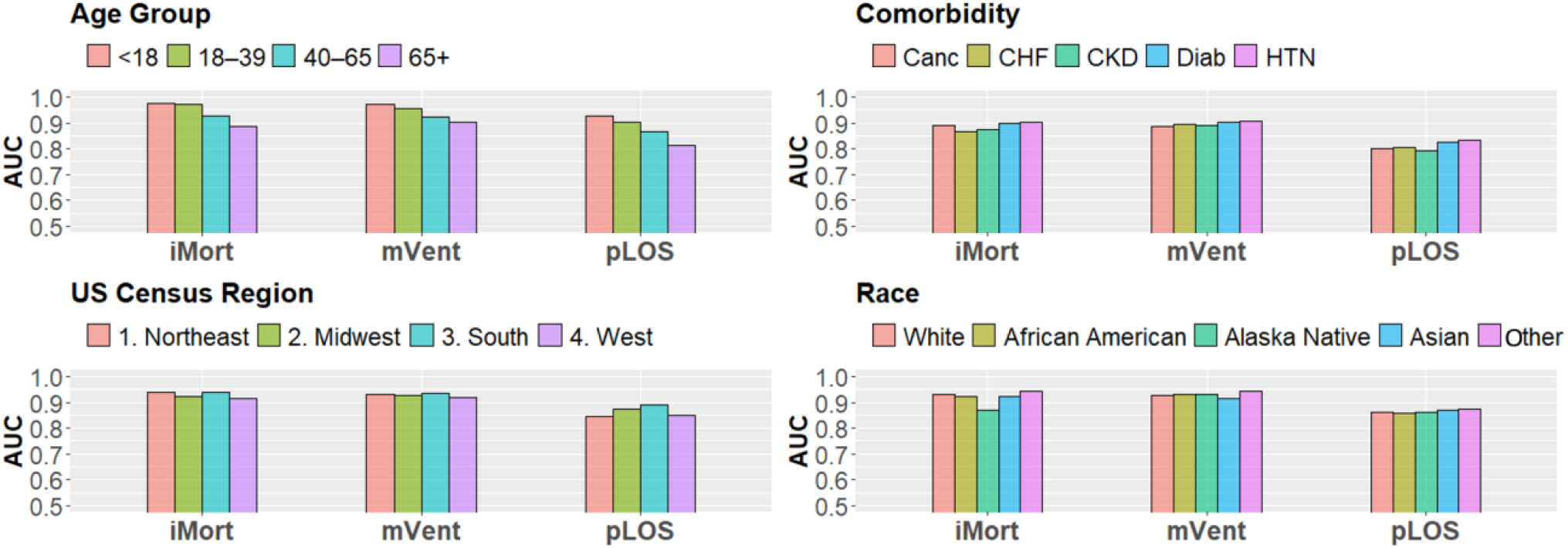
Subgroup analysis using the CRWD multi-hospital test set.

## DISCUSSION

Our experiments showed that CovRNN models trained on a large heterogeneous dataset of approximately 200,000 COVID-19 patients required minimum data curation to achieve high prediction accuracy (AUROC: 97–86%) for different patient clinical outcomes, namely iMort, mVent, and pLOS. CovRNN not only showed high prediction accuracy but also demonstrated good transferability between two large de-identified EHR databases with different structures, good external validity, proper model calibration, and utility of fine-tuning for continuous improvement. In addition, we used integrated gradients as a utility to expose the factors that contribute to the model predicted scores.

CovRNN models were predominant, with consistently higher performance as compared to other baseline methods. Interestingly, we found that the maximum difference between LR, LGBM, and CovRNN models’ AUROC for iMort and mVent was around 3%, whereas it exceeds 6% for pLOS. Similarly, we observed that the pLOS prediction accuracy was highly affected by the inclusion of patient history. Therefore, we can conclude that considering the sequence of events that occurred in the past is of higher importance for the pLOS prediction task versus the iMort and mVent prediction tasks, for which the most recent events are of higher importance.

Although several studies reported predictive machine learning models with prediction accuracy comparable to our,^6,7,23–25^ our model was trained and evaluated on larger, multicenter cohorts from two large, well-known de-identified EHR databases in the United States (a total of more than 300,000 patients). The N3C study^7^ had a similar number of COVID-19 patients (160,000) included in their training set; however, their reported prediction accuracy (AUROC) against iMort and mVent (combined as severity indicator) was only 86%. In addition, the majority of published studies with machine learning models predict the outcome in a very short follow-up window, such as 1 hour to 1 day from the index time-point.^6,25^ Further, some studies did not specify the time window of prediction or used limited historical data.^26^ As the window periods become shorter, the prediction task is easier, and, thus, the accuracy is higher, but the results are less valuable, as physicians can predict those clinical outcomes better without using models. We reported CovRNN survival model results to demonstrate the flexibility of our approach. We believe, however, that predicting the probability of the adverse events’ occurring within the hospital stay should be informative enough for clinicians to make appropriate decisions on admission and may not be limited to a specific time range. Therefore, we also focused on the evaluation and calibration of the binary classification models.

In our study, we included data available on or before the index admission to predict clinical outcomes throughout the hospital stay. Of note, our cohorts also included patients who stayed in observation units without hospitalization. Many physicians often encounter significant dilemmas when deciding the patient’s disposition, such as discharge or higher level of care, in the observation unit or at the time of hospitalization. Further, patients with COVID-19 often progress rapidly, especially after about 7 days from the onset of symptoms, even when they initially present with mild symptoms.^27^ This characteristic clinical course in patients with COVID-19 makes it substantially difficult for clinicians to predict future outcomes on the first day of hospital encounters. Our model is particularly helpful in those clinical scenarios, as the trajectory of the most important clinical outcomes, such as in-hospital mortality, was predicted with specificity of 71% at 95% sensitivity (Supplementary Table 3). As indicated in Supplementary Table 3, the threshold can be easily adjusted to prioritize the sensitivity or specificity to meet the clinicians’ needs. For example, in the situation in which our model predicts the patient’s death with high specificity, physicians could initiate an early discussion of poor outcomes with the patient and/or goals of care in appropriate cases.

The minimal need for data curation and reliance on the power of the deep learning model architecture for learning proper feature representations from large data are key advantages of our CovRNN model. We were able to transfer the model between two completely different datasets that have some differences, particularly in clinical codes distributions. With a simple model fine-tuning step on sample data from the destination dataset, the model consistently achieved high prediction accuracy. Although we focused on COVID-19 patient outcomes, this is a good example for a proof of concept, and we can apply the same methodology to predict different clinical circumstances.

Our study has several limitations. First, our data analysis includes only retrospective data. Despite our efforts to avoid potential bias by separating training, validation, and test datasets as well as external validation on a different data source, potential biases are inevitable. A prospective validation study is warranted, ideally, in hospitals that did not participate in data sharing with the database that we used to secure the validation of transferability. Second, our model focused only on the admission data to predict in-hospital clinical outcomes. It is possible to use multiple time points during the hospital stay to update models to achieve “real-time” predictions. Because minimal data preprocessing is required, our model can be easily modified to use different data points to predict future clinical outcomes.

Third, we performed only a preliminary evaluation for the model predictions explanations, whereby we extracted data from 20 random sample patients and presented their predicted risk scores as well as the contribution score assigned for each medical event and asked infectious disease specialists to evaluate its relevance. Although we acknowledge that this is not a rigorous evaluation method, it demonstrated that our proposed model provides the tool that allows model transparency and helps to engage clinicians and facilitate their judgment on the model predictions. Future work is warranted to systematically evaluate the model’s transparency. Fourth, the dynamics of COVID-19 management in hospitals and patient surges from pandemics have changed over time, which would modify the patient outcomes over time. Thus, the accuracy of our model may be affected in future datasets. For example, the patients who received COVID-19 vaccines likely have different clinical outcomes.^28^ Because our model is trained on historical data, the model can be easily fine-tuned on more current data to improve its prediction accuracy, which is one of the major advantages of deep learning models.

## CONCLUSION

Through benchmarking, we found that our CovRNN can provide accurate and transferable predictive models for a wide range of outcomes and that we can continuously improve upon the model through periodic fine-tuning. Further, our data preparation pipeline is kept to a minimum to facilitate the transferability of the models and facilitate further validation on new data sources. Our model development framework can be further applied to train and evaluate predictive models for different types of clinical events. For clinicians who are fighting COVID-19 on the front lines, there are two potential actionable contributions of our work. Clinicians can (i) fine-tune our pre-trained model on their local data, regardless of the size, establish utility, and then deploy; and (ii) use our comprehensive model development framework to train a predictive model, using their own data.

## Data Availability

The data that support the findings of this study, the Cerner® Real-World COVID-19 Q3 Dataset and Optum® de-identified COVID-19 Electronic Health Record v. 1015 dataset (2007 - 2020), are available for licensing at Cerner Corporation and Optum, Inc., respectively. Data access may require a data-sharing agreement and may incur data access fees.

## DATA AVAILABILITY

The data that support the findings of this study, the Cerner® Real-World COVID-19 Q3 Dataset and Optum® de-identified COVID-19 Electronic Health Record v. 1015 dataset (2007– 2020), are available for licensing at Cerner Corporation and Optum, Inc., respectively. Data access may require a data sharing agreement and may incur data access fees.

## AUTHORS’ CONTRIBUTION

L.R. and D.Z. conceived the idea. L.R. led the design and the implementation of experiments. L.R., K.P., M.N., and B.K. reviewed the evidence before the study. M.N. contributed to the discussion and the model explanation evaluation. Z.X. contributed to the model explanation. L.R., B.M., and K.P. ran the experiments on the OPTUM data. L.R. and Y.Z. extracted the EHR data. W.Z. added the visualizations. L.R. led the writing. B.K., M.N., H.X., and D.Z. also contributed to the writing. AR assessed the study against TRIPOD and PROBAST standards. H.X. and D.Z. supervised the project. L.R. and D.Z. finalized the manuscript. All co-authors reviewed and approved the manuscript.

## COMPETING INTERESTS

The authors have no competing interests to declare.

## ACKNOWLEDGMENTS

We would like to acknowledge the use of the Cerner Real World® dataset and the assistance provided by the University of Texas Health Science Center in Houston (UTHealth) School of Biomedical Informatics (SBMI) Data Service team and the UTHealth Center of Healthcare Data.

## FUNDING

This work was supported by the Cancer Prevention and Research Institute of Texas (CPRIT) Grant No. RP170668 and the UTHealth Innovation for Cancer Prevention Research Training Program Pre-Doctoral Fellowship (CPRIT Grant No. RP160015 and CPRIT Grant No. RP210042).

## Notes

### Competing Interest Statement

The authors have declared no competing interest.

### Author Declarations

1. Non-abbreviated, full names and affiliations of all Ethics Committees / Institutional Review Boards that ruled on ethics of your study. 2. Decision made, i.e. whether ethical approval was given or waived. The Committee for the Protection of Human Subjects at the University of Texas Health Science Center in Houston reviewed the IRB # HSC-SBMI-20-0836 for the "Analysis of COVID-19 related data in Cerner's HealtheDataLab" project. The committee determined the project to qualify for exempt status according to 45 CFR 46.101(b)

## References

1 Coronavirus disease (COVID-19) – World Health Organization. https://www.who.int/emergencies/diseases/novel-coronavirus-2019 (accessed May 29, 2021).

2 CDC. COVID Data Tracker. 2020; published online March 28. https://covid.cdc.gov/covid-data-tracker (accessed March 28, 2021).

3 Wynants L, Van Calster B, Collins GS, et al. Prediction models for diagnosis and prognosis of covid-19 infection: systematic review and critical appraisal. BMJ 2020; 369: m1328.

4 Sperrin M, Grant SW, Peek N. Prediction models for diagnosis and prognosis in Covid-19. BMJ. 2020; 369: m1464.

5 Leeuwenberg AM, Schuit E. Prediction models for COVID-19 clinical decision making. Lancet Digit Health. 2020; 2: e496–7.

6 Schwab P, Mehrjou A, Parbhoo S, et al. Real-time prediction of COVID-19 related mortality using electronic health records. Nat Commun 2021; 12: 1058.

7 Bennett TD, Moffitt RA, Hajagos JG, et al. The National COVID Cohort Collaborative: Clinical Characterization and Early Severity Prediction. medRxiv 2021; published online Jan 13. DOI:10.1101/2021.01.12.21249511.

8 Rasmy L, Wu Y, Wang N, et al. A study of generalizability of recurrent neural network-based predictive models for heart failure onset risk using a large and heterogeneous EHR data set. J Biomed Inform 2018; 84: 11–6.

9 Xiang Y, Ji H, Zhou Y, et al. Asthma Exacerbation Prediction and Risk Factor Analysis Based on a Time-Sensitive, Attentive Neural Network: Retrospective Cohort Study. J Med Internet Res 2020; 22: e16981.

10 Rasmy L, Zhu J, Li Z, Hao X, Zhi D. Simple Recurrent Neural Networks is all we need for clinical events predictions using EHR data. 2019; published online Aug 27. DOI:10.13140/RG.2.2.13199.51368.

11 Wanyan T, Honarvar H, Jaladanki SK, et al. Contrastive Learning Improves Critical Event Prediction in COVID-19 Patients. ArXiv 2021; published online Jan 11. https://www.ncbi.nlm.nih.gov/pubmed/33442560.

12 Rajkomar A, Oren E, Chen K, et al. Scalable and accurate deep learning with electronic health records. npj Digital Medicine 2018; 1: 1–10.

13 Sundararajan M, Taly A, Yan Q. Axiomatic Attribution for Deep Networks. In: International Conference on Machine Learning. PMLR, 2017: 3319–28.

14 UMLS Metathesaurus - MMSL (Multum) - Synopsis. https://www.nlm.nih.gov/research/umls/sourcereleasedocs/current/MMSL/index.html (accessed May 27, 2021).

15 Rasmy L, Tiryaki F, Zhou Y, et al. Representation of EHR data for predictive modeling: a comparison between UMLS and other terminologies. J Am Med Inform Assoc 2020; 27: 1593–9.

16 Harrell FE, Califf RM, Pryor DB, Lee KL, Rosati RA. Evaluating the Yield of Medical Tests. JAMA 1982; 247: 2543–6.

17 Choi E, Schuetz A, Stewart WF, Sun J. Using recurrent neural network models for early detection of heart failure onset. J Am Med Inform Assoc 2017; 24: 361.

18 Choi E, Bahadori MT, Sun J, Kulas J, Schuetz A, Stewart W. RETAIN: An Interpretable Predictive Model for Healthcare using Reverse Time Attention Mechanism. Adv Neural Inf Process Syst 2016; 29. https://proceedings.neurips.cc/paper/2016/file/231141b34c82aa95e48810a9d1b33a79-Paper.pdf (accessed Sept 2, 2021).

19 Modeling asynchronous event sequences with RNNs. J Biomed Inform 2018; 83: 167–77.

20 sklearn.linear_model.LogisticRegression — scikit-learn 0.24.2 documentation. https://scikit-learn.org/stable/modules/generated/sklearn.linear_model.LogisticRegression.html (accessed May 27, 2021).

21 Welcome to LightGBM’s documentation! — LightGBM 3.2.1.99 documentation. https://lightgbm.readthedocs.io/ (accessed May 27, 2021).

22 Katzman JL, Shaham U, Cloninger A, Bates J, Jiang T, Kluger Y. DeepSurv: personalized treatment recommender system using a Cox proportional hazards deep neural network. BMC Med Res Methodol 2018; 18: 1–12.

23 Villegas M, Gonzalez-Agirre A, Gutiérrez-Fandiño A, et al. Predicting the Evolution of COVID-19 Mortality Risk: a Recurrent Neural Network Approach. DOI:10.1101/2020.12.22.20244061.

24 Razavian N, Major VJ, Sudarshan M, et al. A validated, real-time prediction model for favorable outcomes in hospitalized COVID-19 patients. NPJ Digit Med 2020; 3: 130.

25 Yadaw AS, Li Y-C, Bose S, Iyengar R, Bunyavanich S, Pandey G. Clinical features of COVID-19 mortality: development and validation of a clinical prediction model. Lancet Digit Health 2020; 2: e516–25.

26 Estiri H, Strasser ZH, Murphy SN. Individualized prediction of COVID-19 adverse outcomes with MLHO. Sci Rep 2021; 11: 5322.

27 CDC. Healthcare Workers. 2021; published online May 27. https://www.cdc.gov/coronavirus/2019-ncov/hcp/clinical-guidance-management-patients.html (accessed June 7, 2021).

28 COVID-19 Vaccine Breakthrough Infections Reported to CDC — United States, January 1–April 30, 2021. MMWR Morb Mortal Wkly Rep 2021; 70. DOI:10.15585/mmwr.mm7021e3.

