## Supplementary Material A for "CovRNN—A recurrent neural network model for predicting outcomes of COVID-19 patients: model development and validation using EHR data"

**Supplemental Material A: Data Sources**

**Cerner Real World Data^TM^ (CRWD)**

Cerner Real-World Data^TM^ is a national, de-identified, person-centric dataset solution provided by Cerner Corporation to enable researchers to leverage longitudinal electronic health records (EHR) data from contributing organizations. Cerner offers one-year free access to a COVID-19 data science workspace, which includes access to a CRWD COVID-19 de-identified data cohort hosted on the HealtheDataLab^TM^—the Cerner data science ecosystem, built and deployed on Amazon Web Services (AWS).

Data in the CRWD are extracted from the EHR of hospitals and clinics that have consented to such use. Encounters may include pharmacy, clinical, and microbiology laboratory, admission, and billing information from affiliated patient care locations. All admissions, medication orders, dispensing, laboratory orders, and specimens are date- and time-stamped, providing a temporal relationship between treatment patterns and clinical information. Cerner de-identifies the CRWD in compliance with the Health Insurance Portability and Accountability Act (HIPAA).

In our study, we used the latest freely available version (Q3), which includes patient data up to the end of September 2020.

**Optum® de-identified COVID-19 Electronic Health Record** **Dataset (OPTUM)**

Given the urgent need to clinically understand the novel virus of COVID 19, Optum developed a low latency data pipeline that enables minimal data lag, while preserving as much clinical data as possible. The data are sourced from Optum’s longitudinal EHR repository, which is derived from dozens of healthcare provider organizations in the United States, including more than 700 hospitals and 7,000 clinics. The data are certified as de-identified by an independent statistical expert, following HIPAA statistical de-identification rules, and managed according to Optum® customer data-use agreements. The COVID-19 data asset incorporates a wide swath of raw clinical data, including new, unmapped COVID-specific clinical data points from inpatient and ambulatory electronic medical records (EMRs), practice management systems, and numerous other internal systems. Information is processed from across the continuum of care, including acute inpatient stays and outpatient visits. The COVID-19 data capture point of care diagnostics specific to the COVID-19 patient during initial presentation, acute illness, and convalescence, with over 500 mapped labs and bedside observations, including COVID-19 specific testing.

The Optum COVID-19 data elements include patient-level information: demographics, mortality, and clinical interventions, such as medications prescribed and administered. The data are composed of multiple tables that can be linked by a common patient identifier (an anonymous, randomized string of characters). The COVID-19 patient base includes patients in the EHR database who have documented clinical care from January 2007 to the most current monthly data release (October 2020) with a documented diagnosis of COVID-19 or acute respiratory illness after February 1, 2020, and/or documented COVID-19 testing (positive or negative result).

In our study, we used the 1015 version, which includes patient data until October 15, 2020. We only included COVID-19 patients with a hospital stay longer than one day and a confirmed COVID-19 diagnosis, either through positive COVID-19 testing results or a documented COVID-19 diagnosis code (U071).

**Major differences and similarities between CRWD and OPTUM**

A key difference between both datasets is that they applied different de-identification strategies. The CRWD applied date shifting for date de-identification, whereas OPTUM did not apply date shifting for events but, rather, masked the exact day for patient identifiable dates, such as date of birth. OPTUM did not provide the exact encounter disposition for expired patients, whereas this is available on CRWD.

CRWD and OPTUM used nearly the same standard codes for diagnosis, procedures, laboratory, and assessment results; thus, we did not need to apply any terminology normalization. Cerner used Multum codes for medications, whereas OPTUM used NDC codes. Thus, we used the Cerner Multum drug database mapping tool to map the OPTUM medication NDC codes to Multum codes.

The CRWD results table includes the clinical interpretations of laboratory results and does not include the normal range for numerical values. Although the OPTUM laboratory results include the normal range for numerical values, they do not include the interpretation. As in our study, we converted all numerical laboratory results into a “below normal low, normal, or above normal high” classification, by using the interpretation value provided in the CRWD, or by defining the result categories, based on the assigned normal result ranges for OPTUM. In regard to demographics, the OPTUM version that we used did not include a race group for native Alaskans, and the South Atlantic and West South regions were merged, which is not the case in the CRWD. Supplementary Figure 1 shows the geographical coverage for the CRWD and OPTUM cohorts.

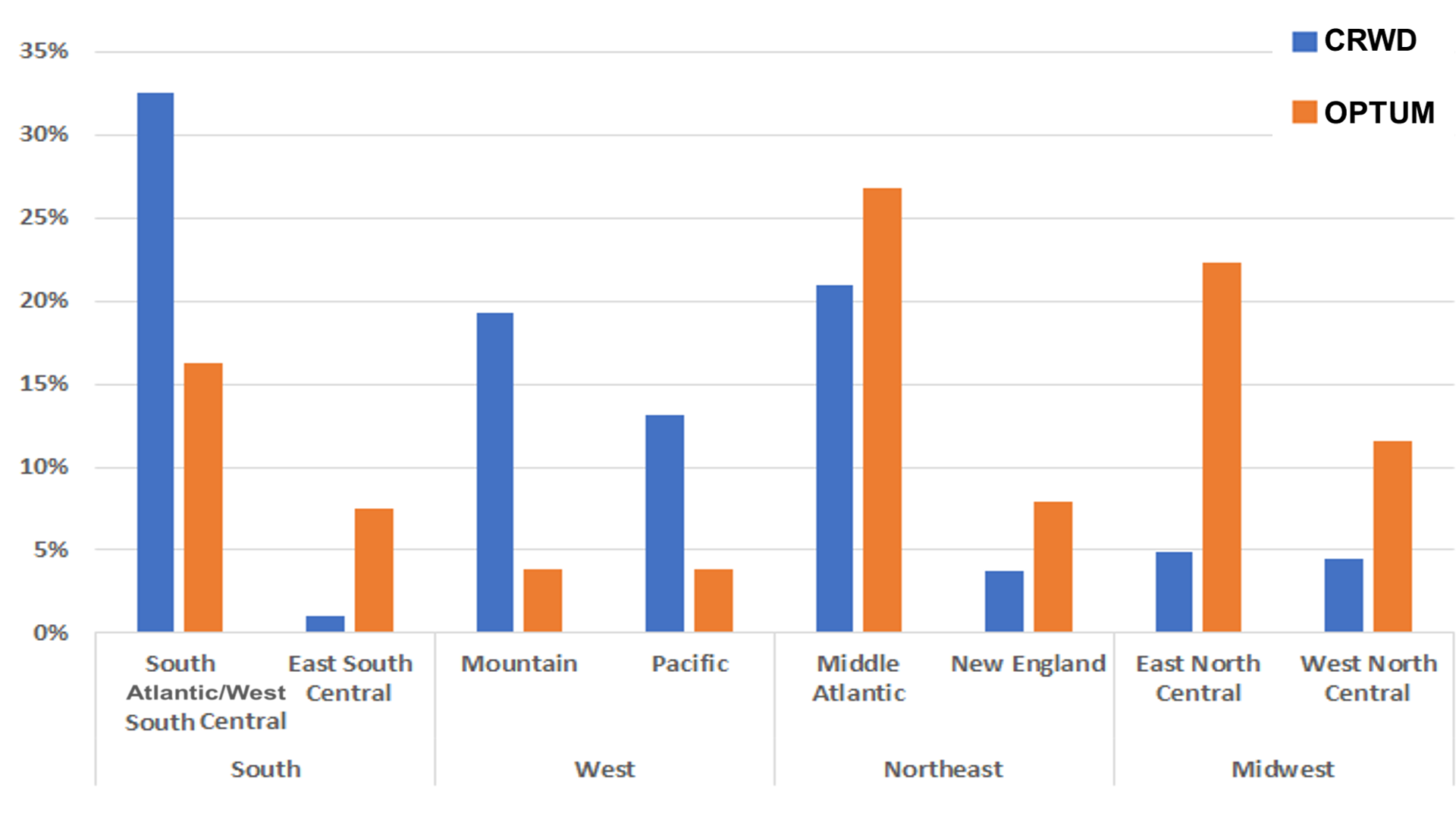

**Supplementary Figure 1. Geographical distribution of CRWD and OPTUM cohorts**
