## Supplementary Table 1 for "CovRNN—A recurrent neural network model for predicting outcomes of COVID-19 patients: model development and validation using EHR data"

**Supplemental Material B: Data Preparation**

We predefined our prediction point as the first COVID-19 admission date, and we refer to it as the index date. For training and internal validation, we excluded all patients who stayed in the hospital for less than 1 day or died within 1 day (24 hrs) to ensure that there was no information leakage and to train the model on more difficult cases. We extracted all patient data available on or before the index date.

For diagnoses, we included the diagnosis code types along with the diagnosis codes recorded before the index admission date. The CRWD and OPTUM datasets used mainly ICD-9, ICD-10, or SNOMED CT codes for recording diagnosis information. Therefore, we relied on these codes without any further normalization. We excluded diagnosis codes of other or unknown types. For medications, as the CRWD used Multum codes, whereas OPTUM used NDC codes, and given that we had access to the NDC to Multum mappings, we converted NDC codes to Multum drug identifiers that correspond to the drug generic name and major dosage form and used the multum drug identifiers and the multum therapeutic categories in our input variables. For procedures, we included all procedure codes, specifically ICD-9PCS, ICD10PCS, CPT, and HCPCS. For laboratory results, in addition to converting to categorical variables, we also mainly used LOINC codes. For an example patient record, we converted the information to look as follows: [ICD9_789.22, loinc_1244-1$High, Multumdnum_d03807, MCat_Antidiabetic, g_Female, r_White, a_87 . . . ].

**Supplementary Table 1: Clinical codes used to define mVent outcome on CRWD**

| **Code Type** | **Codes** |
| --- | --- |
| ICD-10-PCS | 5A1955Z, 5A1945Z, 5A1935Z, 5A09357, 5A09457, 5A09557, 5A09358, 5A0935Z, 5A0945Z, 5A0955Z, 5A09458, 5A09558, 5A09559, 5A09459 |
| ICD-9-PCS | 93.9, 96.71, 96.72 |
| CPT-4 | 94002, 94660, 94003, 78582 |
| SNOMED CT | 47545007, 243142003, 251901004, 26261000175105 |
| LOINC | 19834-1, 19835-8, 19839-0, 19840-8, 19932-3, 19976-0, 19994-3, 19996-8, 20054-3, 20055-0, 20056-8, 20058-4, 20063-4, 20068-3, 20077-4, 20079-0, 20112-9, 20116-0, 20124-4 ,33429-2, 33438-3, 33446-6, 60794-5, 76007-4, 76222-9 |
