## Supplementary Material C for "CovRNN—A recurrent neural network model for predicting outcomes of COVID-19 patients: model development and validation using EHR data"

**Supplemental Material C: Implementation Details**

We used Scikit-learn package v.0.24 for LR, LGBM package v.3.1.1 for LGBM, and Pytorch v.1.7 for CovRNN. For survival evaluation and visualizations, we used lifelines package v.0.23.7. For hyperparameter tuning, we used the Tree-structured Parzen Estimator (TPE) algorithm available through Optuna package v.2.5 to search for the best hyperparameters combination, using a sample cohort extracted from OPTUM data. We evaluated the same on CRWD, and it showed improved model performance compared to the default parameters used in Experiment 1 (Table 4). Therefore, for later results, we used the following hyperparameter: For LGBM, we used a learning rate (lr) of 0.05, feature fraction (proportion of features included on each iteration) as 0.55, min_child_samples (minimum number of data in one leaf) as 77, min_split_gain (minimal gain to perform split) as 0·1, n_estimators (number of boosting iterations) as 150, bagging_fraction (randomly selecting part of the data without resampling) as 0·68, bagging_freq (frequency for bagging) as 4, num_leaves (max number of leaves in one tree) as 139, reg_alpha (L1 regularization) as 0·52. For LR, we found that a weighted L2 regularized model trained with a liblinear solver (algorithm to use in optimization problem) and with C (inverse of regularization strength) as 0·004, was associated with the best performance. For RNN based models, we used the Adagrad optimizer with a starting learning rate (lr) of 0·05, weight decay (L2 penalty) of 0·0001, and the eps (term added to the denominator to improve numerical stability) as 1e-4. We used embedding and hidden dimensions of 64, as they were associated with one of the best performances, as well as the efficient model size and running time. We used a training batch size of 128 for binary classification tasks and 256 for survival models.
