## Supplementary Table 2 for "CovRNN—A recurrent neural network model for predicting outcomes of COVID-19 patients: model development and validation using EHR data"

**Supplementary Table 2: Descriptive analysis for different test sets**

| Characteristics | CRWD Training | CRWD Valid | CRWD Multi– Hospital Test | Hospital 1 | Hospital 2 | OPTUM Fine-tuning | OPTUM Test |
| --- | --- | --- | --- | --- | --- | --- | --- |
|  | *n* = 170,626 | *n* = 24,378 | *n* = 48,781 | *n* = 3,469 | *n* = 706 | *n* = 29,416 | *n* = 6,724 |
| Age on index Median (IQR) | 57 (36–72) | 57 (35–72) | 57 (36–72) | 58 (40–71) | 43 (30–57) | 60 (44–72) | 59 (43–71) |
| Gender |  |  |  |  |  |  |  |
| Female | 89,844 (52%) | 12,843 (52%) | 25,693 (52%) | 1,814 (52%) | 346 (49%) | 14,898 (50%) | 3,339 (49%) |
| Male | 80,269 (47%) | 11,467 (47%) | 22,915 (46%) | 1,643 (47%) | 359 (50%) | 14,505 (49%) | 3,380 (50%) |
| Race & Ethnicity |  |  |  |  |  |  |  |
| Caucasian | 116,342 (68%) | 16,577 (68%) | 33,278 (68%) | 1,786 (51%) | 623 (88%) | 16,047 (54%) | 3,657 (54%) |
| African American | 24,748 (14%) | 3,545 (14%) | 7,034 (14%) | 1,395 (40%) | 40 (5%) | 6,427 (21%) | 1,409 (20%) |
| Asian | 3,843 (2%) | 539 (2%) | 1,088 (2%) | 15 (0%) | 9 (1%) | 742 (2%) | 188 (2%) |
| American Indian /Alaska Native | 2,919 (1%) | 453 (1%) | 908 (1%) | 4 (0%) | 1 (0%) | NA | NA |
| Hispanic | 50,114 (29%) | 7,247 (29%) | 14,113 (28%) | 101 (2%) | 493 (69%) | 4,708 (16%) | 1,074 (15%) |
| Comorbidities |  |  |  |  |  |  |  |
| Hypertension (HTN) | 78,260 (45%) | 11,274 (46%) | 22,576 (46%) | 2,105 (60%) | 172 (24%) | 18,039 (61%) | 3,996 (59%) |
| Diabetes (DM) | 43,918 (25%) | 6,326 (25%) | 12,471 (25%) | 1,182 (34%) | 126 (17%) | 10,706 (36%) | 2236 (33%) |
| Congestive Heart Failure (CHF) | 24,598 (14%) | 3,565 (14%) | 7,189 (14%) | 643 (18%) | 45 (6%) | 5,428 (18%) | 1,140 (16%) |
| Chronic Kidney Disease (CKD) | 23,827 (13%) | 3,469 (14%) | 6,794 (13%) | 661 (19%) | 38 (5%) | 6,208 (21%) | 1,309 (19%) |
| Cancer | 13,074 (7%) | 1,910 (7%) | 3,826 (7%) | 310 (8%) | 25 (3%) | 4,229 (14%) | 865 (12%) |
| Outcomes |  |  |  |  |  |  |  |
| Mortality (iMort) | 9,324 (5%) | 1,321 (5%) | 2,666 (5%) | 263 (7%) | 33 (4%) | 3,946 (13%) | 885 (13%) |
| Median TTE | 3 (1–6) | 3 (1–6) | 3 (1–6) | 2 (1–6) | 1 (1–5) | 5 (3–10) | 5 (3–10) |
| Mechanical ventilation (mVent) | 23,127 (13%) | 3,225 (13%) | 6,556 (13%) | 496 (14%) | 101 (14%) | 7,845 (26%) | 1,737 (25%) |
| Intubated on first day (% of mVent) | 12,270 (53%) | 1,703 (52%) | 3,557 (54%) | 215 (43%) | 66 (65%) | 3,676 (46%) | 790 (45%) |
| Median TTE | 2 (1–4) | 2 (1–4) | 2 (1–4) | 2 (1–4) | 1 (1–3) | 3 (2–7) | 3 (2–7) |
| Length of stay Median (IQR) | 3 (1–6) | 3 (1–6) | 3 (1–6) | 2 (1–6) | 1 (1–5) | 5 (3–10) | 5 (3–10) |
