## Supplementary Table 3 for "CovRNN—A recurrent neural network model for predicting outcomes of COVID-19 patients: model development and validation using EHR data"

**Supplementary Table 3: Additional prediction accuracy metrics for CovRNN binary classification models**

| **Cohort** | **task** | **AUROC** | **AUPRC** | **Specificity @ 95% Sensitivity** | **Sensitivity*** | **F1-score*** | **Specificity*** |
| --- | --- | --- | --- | --- | --- | --- | --- |
| CRWD Training Set | iMort | 95·33% | 63·74% | 79·62% | 90·35% | 42·02% | 86·15% |
|  | mVent | 95·91% | 86·36% | 76·88% | 89·95% | 65·41% | 86·66% |
|  | pLOS | 90·75% | 71·04% | 64·98% | 86·70% | 61·38% | 77·90% |
| CRWD Valid Set | iMort | 92·30% | 49·75% | 67·52% | 83·80% | 38·38% | 85·51% |
|  | mVent | 92·56% | 77·93% | 60·88% | 83·01% | 59·33% | 85·24% |
|  | pLOS | 86·58% | 59·07% | 56·43% | 80·32% | 55·83% | 75·70% |
| CRWD Multi-hospital Test Set | iMort | 93·03% | 52·84% | 70·93% | 84·92% | 39·22% | 85·65% |
|  | mVent | 92·90% | 79·51% | 63·48% | 83·39% | 59·73% | 85·12% |
|  | pLOS | 86·50% | 60·00% | 55·56% | 79·74% | 56·51% | 76·28% |
| Hospital 1 | iMort | 91·77% | 51·24% | 70·87% | 82·51% | 44·24% | 84·37% |
|  | mVent | 91·54% | 73·71% | 62·33% | 76·01% | 61·20% | 87·92% |
|  | pLOS | 87·15% | 57·86% | 59·68% | 76·96% | 58·53% | 79·64% |
| Hospital 2 | iMort | 97·00% | 59·57% | 86·18% | 90·91% | 51·72% | 92·12% |
|  | mVent | 96·02% | 85·25% | 85·12% | 83·17% | 69·42% | 90·58% |
|  | pLOS | 88·33% | 61·41% | 63·95% | 75·42% | 55·80% | 80·95% |
| OPTUM Test Set | iMort | 91·27% | 70·63% | 59·34% | 82·94% | 56·44% | 83·18% |
|  | mVent | 91·46% | 83·19% | 55·02% | 89·06% | 67·54% | 73·99% |
|  | pLOS | 80·97% | 70·24% | 35·52% | 85·97% | 64·42% | 57·25% |

******Sensitivity, Specificity, and F1-Score are at the best identified threshold of 7·5% for iMort, 10% for mVent, and 20% for pLOS*
