## Supplementary Material E for "CovRNN—A recurrent neural network model for predicting outcomes of COVID-19 patients: model development and validation using EHR data"

**PROBAST**

(Prediction model study Risk Of Bias Assessment Tool)

Published in Annals of Internal Medicine (freely available):

1. [PROBAST: A Tool to Assess the Risk of Bias and Applicability of Prediction Model Studies](https://annals.org/aim/fullarticle/2719961/probast-tool-assess-risk-bias-applicability-prediction-model-studies)
2. [PROBAST: A Tool to Assess Risk of Bias and Applicability of Prediction Model Studies: Explanation](https://annals.org/aim/fullarticle/2719962/probast-tool-assess-risk-bias-applicability-prediction-model-studies-explanation) [and Elaboration](https://annals.org/aim/fullarticle/2719962/probast-tool-assess-risk-bias-applicability-prediction-model-studies-explanation)

### What does PROBAST assess?

PROBAST assesses both the *risk of bias* and *concerns regarding applicability* of a study that evaluates (develops, validates or updates) a multivariable diagnostic or prognostic prediction model. It is designed to assess primary studies included in a systematic review.

*Bias* occurs if systematic flaws or limitations in the design, conduct or analysis of a primary study distort the results. For the purpose of prediction modelling studies, we have defined *risk of bias* to occur when shortcomings in the study design, conduct or analysis lead to systematically distorted estimates of a model’s predictive performance or to an inadequate model to address the research question. Model predictive performance is typically evaluated using calibration, discrimination and sometimes classification measures, and these are likely inaccurately estimated in studies with high risk of bias. *Applicability* refers to the extent to which the prediction model from the primary study matches your systematic review question, for example in terms of the participants, predictors or outcome of interest.

A primary study may include the development and/or validation or update of more than one prediction model. A PROBAST assessment should be completed for each distinct model that is developed, validated or updated (extended) for making individualised predictions. Where a publication assesses multiple prediction models, only complete a PROBAST assessment for those models that meet the inclusion criteria for your systematic review. Please note that subsequent use of the term “model” includes derivatives of models, such as simplified risk scores, nomograms, or recalibrations of models.

PROBAST is not designed for all multivariable diagnostic or prognostic studies. For example, studies using multivariable models to identify predictors associated with an outcome but not attempting to develop a model for making individualised predictions are not covered by PROBAST.

PROBAST includes four steps.

|  | **Step** | **Task** | **When to complete** |
| --- | --- | --- | --- |
|  | **1** | Specify your systematic review question(s) | Once per systematic review |
|  | **2** | Classify the type of prediction model evaluation | Once for each model of interest in each publication being assessed, for each relevant outcome |
|  | **3** | Assess risk of bias and applicability | Once for each development and validation of each distinct prediction model in a publication |
|  | **4** | Overall judgment | Once for each development and validation of each distinct prediction model in a publication |

If this is your first time using PROBAST, we strongly recommend reading the detailed explanation and elaboration (E&E, see link above) paper and to check the examples on [www.probast.org](http://www.probast.org/)

### Step 1: Specify your systematic review question

State your systematic review question to facilitate the assessment of the applicability of the evaluated models to your question. *The following table should be completed once per systematic review.*

| **Criteria** | **Specify your systematic review question** |
| --- | --- |
| *Intended use of model:* | A time-aware deep learning model to predict COVID-19 patients outcomes that require minimal data preprocessing. |
| ***Participants*** *including selection criteria and setting:* | We extracted our main training cohort from Cerner Real-world (CRWD) COVID-19 Q3 cohort which included information for COVID-19 patients from 87 health systems until the end of September 2020. Eligible patients had a minimum of one emergency or inpatient encounter with a diagnosis code that could be associated with COVID-19 exposure or infection, or a positive result for a COVID-19 laboratory test. For our study, we excluded all patients who have less than one day of information after their first COVID-19 admission as well as patients who had confusing dates like discharge dates before the hospitalization start date. Our cohort included 247,960 patients, from which we excluded two hospitals data to use for external validation and the remaining subjects were split into training, validation, and test sets with the ratio of 7:1:2. Therefore, all our reported prediction accuracy metrics on CRWD was the results on a held out test set of 48,781 patients from around 85 health systems. For external validation, we evaluated the model on two randomly selected held out hospitals from CRWD, Hospital 1 from the south region with 3,469 patients and Hospital 2 from the west region with 706 patients. For further external validation we extracted a cohort of 36,140 patients from the OPTUM (COVID-19 v.1015) dataset. |
| ***Predictors*** *(used in prediction modelling), including types of predictors (e.g. history, clinical examination, biochemical markers, imaging tests), time of measurement, specific measurement issues (e.g., any requirements/ prohibitions for specialized equipment):* | We extracted all patient information on or before the date of their first hospital admission with COVID-19, including demographics, diagnosis, medication, procedures, laboratory results, and observations. We utilized standard terminologies in common use like ICD 9, ICD10, SNOMED CT, LOINC, Multum codes for medications, CPT-4, HCPCS, ICD-9 PCs, and ICD-10 PCs for procedures. Such standard terminologies are readily available in the majority of EHR systems for interoperability facilitation. In cases where a coding system is not used such as Multum codes for medication, pre-existing mapping tools are available13.  The majority of our features were categorical such as diagnosis, medications, and procedures. For numeric variables like laboratory results and age we convert those to categorical variables as well. |
| *Outcome to be predicted:* | Our prediction tasks include the prediction of COVID-19 patients’ in-hospital mortality (iMort), need for mechanical ventilation (mVent), and prolonged length of stay (pLOS), on admission. For iMort event definition, we relied on the preassigned flags on CRWD along with the encounter discharge disposition. The iMort event definition was slightly different on the OPTUM data and there were no clear discharge disposition indicating patient in-hospital death, rather we used the date of death and compare against the hospitalization discharge date to assign the proper label. For mVent, we mainly used all relevant mechanical ventilation procedure codes to define the outcome. Additionally on CRWD, we used other relevant observations or laboratory results not only to identify the instant of the event, but also to identify the earliest time of the event. For both iMort and mVent prediction tasks, we trained both survival and binary classification based prediction models. We defined pLOS as a binary indicator for hospitalizations longer than 7 days, as the median length of stay (LOS) in both CRWD and OPTUM cohorts were 3 and 5 respectively, and we only trained a binary classification model for the pLOS task.  For Binary classification tasks we compared our proposed GRU based model, against machine learning algorithms like Logistic regression (LR)15 and light gradient boost machine (LGBM)16.  For survival prediction, we utilized the DeepSurv17 architecture, while replacing the multiple layer perceptrons (MLP) with GRU for better sequential information modeling. We were unable to adequately compare against machine learning survival models like random survival forest (RSF) for computational resource issues especially with the increased number of covariates and large training set size. |

### Step 2: Classify the type of prediction model evaluation

Use the following table to classify the evaluation as model development, model validation or model update, or combination. Different signalling questions apply for different types of prediction model evaluation. If the evaluation does not fit one of these classifications then PROBAST should not be used.

| **Classify the evaluation based on its aim** | | | |
| --- | --- | --- | --- |
| **Type of prediction study** | **PROBAST boxes to complete** | **Tick as appropriate** | **Definition for type of prediction model study** |
| Development only | Development |  | Prediction model development without external validation. These studies may include internal validation methods, such as bootstrapping and cross-validation techniques. |
| Development and validation | Development and validation | X | Prediction model development combined with external validation in other participants in the same article. |
| Validation only | Validation |  | External validation of existing (previously developed) model in other participants. |

| *This table should be completed once for each publication being assessed and for each relevant outcome in your review.* |
| --- |
| **Publication reference** |
| **Models of interest** |
| **Outcome of interest** |

### Step 3: Assess risk of bias and applicability

PROBAST is structured as four key domains. Each domain is judged for risk of bias (low, high or unclear) and includes signalling questions to help make judgements. Signalling questions are rated as yes (Y), probably yes (PY), probably no (PN), no (N) or no information (NI). All signalling questions are phrased so that “yes” indicates absence of bias. Any signalling question rated as “no” or “probably no” flags the potential for bias; you will need to use your judgement to determine whether the domain should be rated as “high”, “low” or “unclear” risk of bias. The guidance document contains further instructions and examples on rating signalling questions and risk of bias for each domain.

The first three domains are also rated for concerns regarding applicability (low/ high/ unclear) to your review question defined above.

*Complete all domains separately for each evaluation of a distinct model. Shaded boxes indicate where signalling questions do not apply and should not be answered.*

| **DOMAIN 1: Participants** | |
| --- | --- |
| **A. Risk of Bias** | |
| *Describe the sources of data and criteria for participant selection:*  We extracted our main training cohort from Cerner Real-world (CRWD) COVID-19 Q3 cohort which included information for COVID-19 patients from 87 health systems until the end of September 2020. Eligible patients had a minimum of one emergency or inpatient encounter with a diagnosis code that could be associated with COVID-19 exposure or infection, or a positive result for a COVID-19 laboratory test. Our cohort included 247,960 patients, from which we excluded two hospitals data to use for external validation and the remaining subjects were split into training, validation, and test sets with the ratio of 7:1:2. For further external validation we extracted a cohort of 36,140 patients from the OPTUM (COVID-19 v.1015) dataset. | |
|  | Dev Val |
| 1.1 Were appropriate data sources used, e.g. cohort, RCT or nested case-contro data? Yes. | l study X X |
| 1.2 Were all inclusions and exclusions of participants appropriate? Yes |  |
| **Risk of bias introduced by selection of participants RISK:**  *(low/ high/ un* | Low  *clear)* |
| *Rationale of bias rating:*  The data analysis only includes retrospective data and the model only focused on the admission data to predict in-hospital clinical outcomes. The study uses appropriate means to prevent bias, measure outcomes, and analyze and report results. | |
| **B. Applicability** | |
| *Describe included participants, setting and dates:*  We extracted our main training cohort from Cerner Real-world (CRWD) COVID-19 Q3 cohort which included information for COVID-19 patients from 87 health systems until the end of September 2020. | |
| **Concern that the included participants and setting do not match CONCERN the review question** *(low/ high/ un* | **: Low**  *clear)* |
| *Rationale of applicability rating:*  In this study, our main objective was to develop an accurate predictive model for COVID-19 patients outcomes at the time of admission, that can be implemented easily with minimal data-preprocessing requirements. The retrospective COVID data utilized matches the research question. | |

| **DOMAIN 2: Predictors** | |
| --- | --- |
| **A. Risk of Bias** | |
| *List and describe predictors included in the final model, e.g. definition and timing of assessment:*  we propose a time-aware deep learning model to predict COVID-19 patients outcomes that require minimal data preprocessing. We used patient data available within the EHR on and before the day of admission and trained recurrent neural network (RNN) models to predict the future risk of three outcomes: in-hospital mortality, need for mechanical ventilation, and long length of stay (LOS >7 days). Our models were developed and validated using a 247,960 patients cohort from 87 health systems from Cerner Real World Dataset (CRWD). The transferability of our model was validated using 36,140 patients from OPTUM. | |
|  | Dev Val |
| 2.1 Were predictors defined and assessed in a similar way for all participants? | Y Y Yes |
| 2.2 Were predictor assessments made without knowledge of outcome data? | Y Y Yes |
| 2.3 Are all predictors available at the time the model is intended to be used? | Y Y Yes |
| **Risk of bias introduced by predictors or their assessment RISK:**  *(low/ high/ un* | *clear) Low* |
| *Rationale of bias rating:*  *Used a valid approach utilizing retrospective data, and means to prevent bias, measure outcomes, and analyze and report results.*  *O*ur proposed model was based on a gated type of recurrent neural networks (RNN), a sequential deep learning architecture, namely gated recurrent unit (GRU). Our model was designed to consume all diagnoses, medications, laboratory results, and other clinical events information readily available in the EHR before or on the index date to predict the patient outcomes, without any need for specific feature selection or missing values imputation, for convenience and practicality. Our proposed model considered the temporal nature of patient history and gave more weight for most recent events compared to distant events that happened years ago. Furthermore, for iMort and mVent prediction tasks, we trained both survival and binary classification models, as our framework allows that to fit different clinical needs for healthcare workers confronting COVID-19. | |
| **B. Applicability** | |
| Concern that the definition, assessment or timing of predictors in **CONCERN**  the model do not match the review question *(low/ high/ un* | **:**  *clear) Low* |
| *Rationale of applicability rating:*  Predicting patient outcomes in patients with COVID-19 in an early stage is a critical need. | |

| **DOMAIN 3: Outcome** | |
| --- | --- |
| **A. Risk of Bias** | |
| *Describe the outcome, how it was defined and determined, and the time interval between predictor assessment and outcome determination:*  Our experiments showed that our GRU based models trained on large heterogeneous dataset of around 200,000 COVID-19 patients required minimum data curation to achieve high prediction accuracy (AUC: 97-86%) for different patient clinical outcomes, namely iMort, mVent and pLOS. Our models were not only showing high prediction accuracy but it demonstrated good transferability between two large deidentified EHR databases with different structures, good external validity, proper model calibration, and the utility of fine-tuning for continuous improvement. Our model was trained and evaluated on larger multicenter cohorts from two large well-known de-identified EHR databases in the U.S. (A total of more than 300,000 patients) . The study was able to transfer the model between two completely different datasets that have some differences especially in clinical codes distributions. With a simple model fine-tuning step on a sample data from the destination dataset, the model consistently achieved high prediction accuracy. Although in this study we focused on COVID-19 patients outcomes, this is a good example for a proof of concept and we can apply the same methodology to predict different clinical circumstances. | |
|  | Dev Val |
| 3.1 Was the outcome determined appropriately? | Y Yes |
| 3.2 Was a pre-specified or standard outcome definition used? | Y Yes |
| 3.3 Were predictors excluded from the outcome definition? | Y Yes |
| 3.4 Was the outcome defined and determined in a similar way for all participants? Y Yes | |
| 3.5 Was the outcome determined without knowledge of predictor information? | Y Y YYes |
| 3.6 Was the time interval between predictor assessment and outcome determi appropriate? | Nation Yes |
| **Risk of bias introduced by the outcome or its determination RISK: Low**  *(low/ high/ unclear)* | |
| *Rationale of bias rating:*  *Efforts to reduce Bias include separating train, validation, and test datasets as well as external validation on a different data source.* | |
| **B. Applicability** | |
| *At what time point was the outcome determined:*  Our experiments showed that our GRU based models trained on large heterogeneous dataset of around 200,000 COVID-19 patients required minimum data curation to achieve high prediction accuracy (AUC: 97-86%) for different patient clinical outcomes, namely iMort, mVent and pLOS. Our models were not only showing high prediction accuracy but it demonstrated good transferability between two large deidentified EHR databases with different structures, good external validity, proper model calibration, and the utility of fine-tuning for continuous improvement. Additionally, We used integrated gradients to provide clinicians with a utility to understand the model predicted scores. Timing?  *If a composite outcome was used, describe the relative frequency/distribution of each contributing outcome:* | |
| **Concern that the outcome, its definition, timing or CONCERN: Low-determination do not match the review question** *(low/ high/ unclear) LLlLoLow* | |
| *Rationale of applicability rating: The Determination matches the research question.* | |

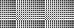

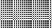

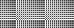

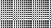

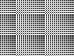

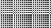

| **DOMAIN 4: Analysis** | |
| --- | --- |
| **Risk of Bias** | |
| *Describe numbers of participants, number of candidate predictors, outcome events and events per candidate predictor:*  *We extracted our main training cohort from Cerner Real-world (CRWD) COVID-19 Q3 cohort which included information for COVID-19 patients from 87 health systems until the end of September 2020. Eligible patients had a minimum of one emergency or inpatient encounter with a diagnosis code that could be associated with COVID-19 exposure or infection, or a positive result for a COVID-19 laboratory test. For our study, we excluded all patients who have less than one day of information after their first COVID-19 admission as well as patients who had confusing dates like discharge dates before the hospitalization start date. Our cohort included 247,960 patients, from which we excluded two hospitals data to use for external validation and the remaining subjects were split into training, validation, and test sets with the ratio of 7:1:2. Therefore, all our reported prediction accuracy metrics on CRWD was the results on a held out test set of 48,781 patients from around 85 health systems. For external validation, we evaluated the model on two randomly selected held out hospitals from CRWD, Hospital 1 from the south region with 3,469 patients and Hospital 2 from the west region with 706 patients. For further external validation we extracted a cohort of 36,140 patients from the OPTUM (COVID-19 v.1015) dataset. Further description of CRWD and OPTUM cohorts, along with differences and commonalities are available in supplementary material A. Table 1 shows the descriptive analysis of both CRWD and OPTUM cohorts.*  We extracted all patient information on or before the date of their first hospital admission with COVID-19, including demographics, diagnosis, medication, procedures, laboratory results, and observations. We utilized standard terminologies in common use like ICD 9, ICD10, SNOMED CT, LOINC, Multum codes for medications, CPT-4, HCPCS, ICD-9 PCs, and ICD-10 PCs for procedures. Such standard terminologies are readily available in the majority of EHR systems for interoperability facilitation. In cases where a coding system is not used such as Multum codes for medication, pre-existing mapping tools are available13.  The majority of our features were categorical such as diagnosis, medications, and procedures. For numeric variables like laboratory results and age we convert those to categorical variables as well. For example, for Laboratory results we used the “below normal low, normal, or above normal high” classification using the interpretation value provided in the CRWD rather than using the actual numerical value, while for OPTUM, we defined the result categories, based on the assigned normal result ranges. By doing so we can further convert our input either to multi-hot or embedding matrices to feed to our models. Based on our previous study14, we decided to use the clinical information in the coding standards it was recorded with, as the normalization of those codes provides minimal gain14. Further details of our data curation is available in Supplementary Material B. Our data curation pipeline will be available in our github repository | |
| *Describe how the model was developed (for example in regards to modelling technique (e.g. survival or logistic modelling), predictor selection, and risk group definition):*  *Our proposed model was based on a gated type of recurrent neural networks (RNN), a sequential deep learning architecture, namely gated recurrent unit (GRU). Our model was designed to consume all diagnoses, medications, laboratory results, and other clinical events information readily available in the EHR before or on the index date to predict the patient outcomes, without any need for specific feature selection or missing values imputation, for convenience and practicality.Our proposed model considered the temporal nature of patient history and gave more weight for most recent events compared to distant events that happened years ago. Furthermore, for iMort and mVent prediction tasks, we trained both survival and binary classification models, as our framework allows that to fit different clinical needs for healthcare workers confronting COVID-19.* | |
| *Describe whether and how the model was validated, either internally (e.g. bootstrapping, cross validation, random split sample) or externally (e.g. temporal validation, geographical validation, different setting, different type of participants):*  *For Binary classification tasks we compared our proposed GRU based model, against machine learning algorithms like Logistic regression (LR)15 and light gradient boost machine (LGBM)16.*  *For survival prediction, we utilized the DeepSurv17 architecture, while replacing the multiple layer perceptrons (MLP) with GRU for better sequential information modeling. We were unable to adequately compare against machine learning survival models like random survival forest (RSF) for computational resource issues especially with the increased number of covariates and large training set size.* | |
| *Describe the performance measures of the model, e.g. (re)calibration, discrimination, (re)classification, net benefit, and whether they were adjusted for optimism:* | |
| *Describe any participants who were excluded from the analysis:* | |
| *Describe missing data on predictors and outcomes as well as methods used for missing data:* | |
|  | Dev Val |
| 4.1 Were there a reasonable number of participants with the outcome? | Y Yes |
| 4.2 Were continuous and categorical predictors handled appropriately? | Y Yes |
| 4.3 Were all enrolled participants included in the analysis? | N No |
| 4.4 Were participants with missing data handled appropriately? | Y Yes |
| 4.5 Was selection of predictors based on univariable analysis avoided? | Y Y |
| 4.6 Were complexities in the data (e.g. censoring, competing risks, sampling of controls) accounted for appropriately? Yes Yes Y Yes | |
| 4.7 Were relevant model performance measures evaluated appropriately? | Yes |
| 4.8 Were model overfitting and optimism in model performance accounted for? | Yes |
| 4.9 Do predictors and their assigned weights in the final model correspond to the re from multivariable analysis? Yes | sults |
| **Risk of bias introduced by the analysis RISK:**  *(low/ high/ un* | *clear) Low* |
| *Rationale of bias rating:*  *The above questions we considered in the study We focused more on the evaluation and calibration of the binary classification models.*  *Our prediction tasks include the prediction of COVID-19 patients’ in-hospital mortality (iMort), need for mechanical ventilation (mVent), and prolonged length of stay (pLOS), on admission. For iMort event definition, we relied on the preassigned flags on CRWD along with the encounter discharge disposition. The iMort event definition was slightly different on the OPTUM data and there were no clear discharge disposition indicating patient in-hospital death, rather we used the date of death and compare against the hospitalization discharge date to assign the proper label. For mVent, we mainly used all relevant mechanical ventilation procedure codes to define the outcome. Additionally on CRWD, we used other relevant observations or laboratory results not only to identify the instant of the event, but also to identify the earliest time of the event. For both iMort and mVent prediction tasks, we trained both survival and binary classification based prediction models. We defined pLOS as a binary indicator for hospitalizations longer than 7 days, as the median length of stay (LOS) in both CRWD and OPTUM cohorts were 3 and 5 respectively, and we only trained a binary classification model for the pLOS task.* | |

### Step 4: Overall assessment

Use the following tables to reach overall judgements about risk of bias and concerns regarding applicability of the prediction model evaluation (development and/or validation) across all assessed domains.

*Complete for each evaluation of a distinct model.*

| **Reaching an overall judgement about risk of bias of the prediction model evaluation** | | |
| --- | --- | --- |
|  | **Low risk of bias** | If all domains were rated low risk of bias.  If a prediction model was developed without any external validation, and it was rated as low risk of bias for all domains, consider downgrading to **high risk of bias**. Such a model can only be considered as low risk of bias, if the development was based on a very large data set and included some form of internal validation. |
|  | **High risk of bias** | If at least one domain is judged to be at **high risk of bias**. |
|  | **Unclear risk of bias** | If an unclear risk of bias was noted in at least one domain and it was low risk for all other domains. |

| **Reaching an overall judgement about applicability of the prediction model evaluation** | |
| --- | --- |
| **Low concerns regarding applicability** | If low concerns regarding applicability for all domains, the prediction model evaluation is judged to have **low concerns regarding applicability**. |
| **High concerns regarding applicability** | If high concerns regarding applicability for at least one domain, the prediction model evaluation is judged to have **high concerns regarding applicability**. |
| **Unclear concerns regarding applicability** | If unclear concerns (but no “high concern”) regarding applicability for at least one domain, the prediction model evaluation is judged to have **unclear concerns regarding applicability** overall. |

| **Overall judgement about risk of bias and applicability of the prediction model evaluation** | |  |
| --- | --- | --- |
| **Overall judgement of risk of bias** | **RISK:**  *(low/ high/ unclear)* | Low |
| *Summary of sources of potential bias:*  The approach and uses appropriate means to prevent bias were utilized. The measurement, analysis and report results we identified | |  |
| **Overall judgement of applicability** | **CONCERN:**  *(low/ high/ unclear)* | High |
| *Summary of applicability concerns:*  Predicting patient outcomes in patients with COVID-19 in an early stage is a critical need. Though multiple machine learning models have been proposed to solve this problem, they have not been validated or implemented outside of the original study site owing to the risk of bias and need for extensive data pre-processing and feature engineering.  Through benchmarking, we found that simple GRU based models can provide accurate and transferable predictive models for a wide range of outcomes, that we can continuously improve upon through periodic fine-tuning. | |  |
