## Supplementary Figure for "CovRNN—A recurrent neural network model for predicting outcomes of COVID-19 patients: model development and validation using EHR data"

**Supplemental Material F: Additional Figures**

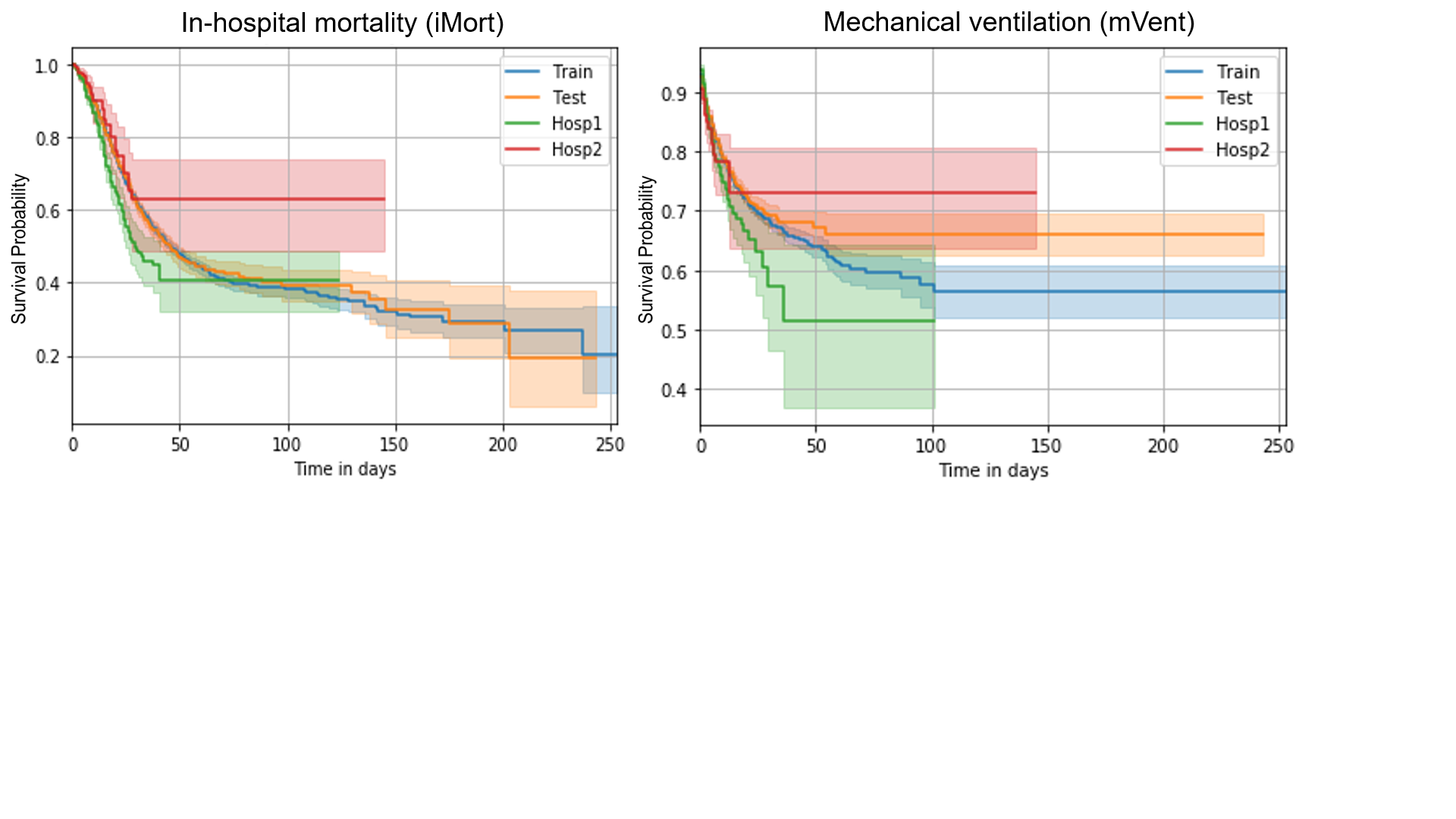

**Supplementary Figure 2. K-M curve of in-hospital mortality and mechanical ventilation CRWD cohorts.**

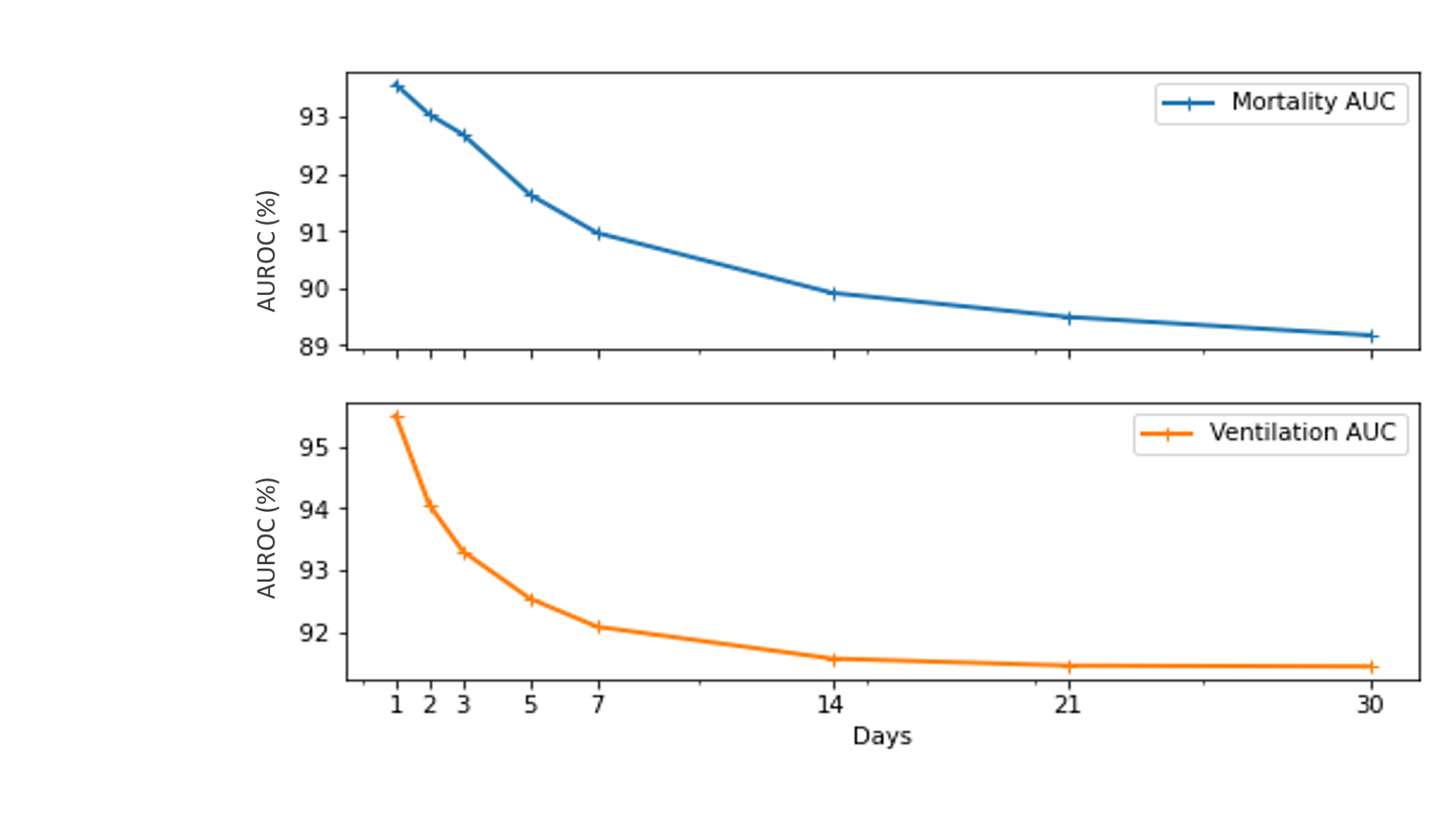

**Supplementary Figure 3. AUROC across different time windows, using iMORT-Surv and mVent-Surv on the CRWD multi-hospital test set.**

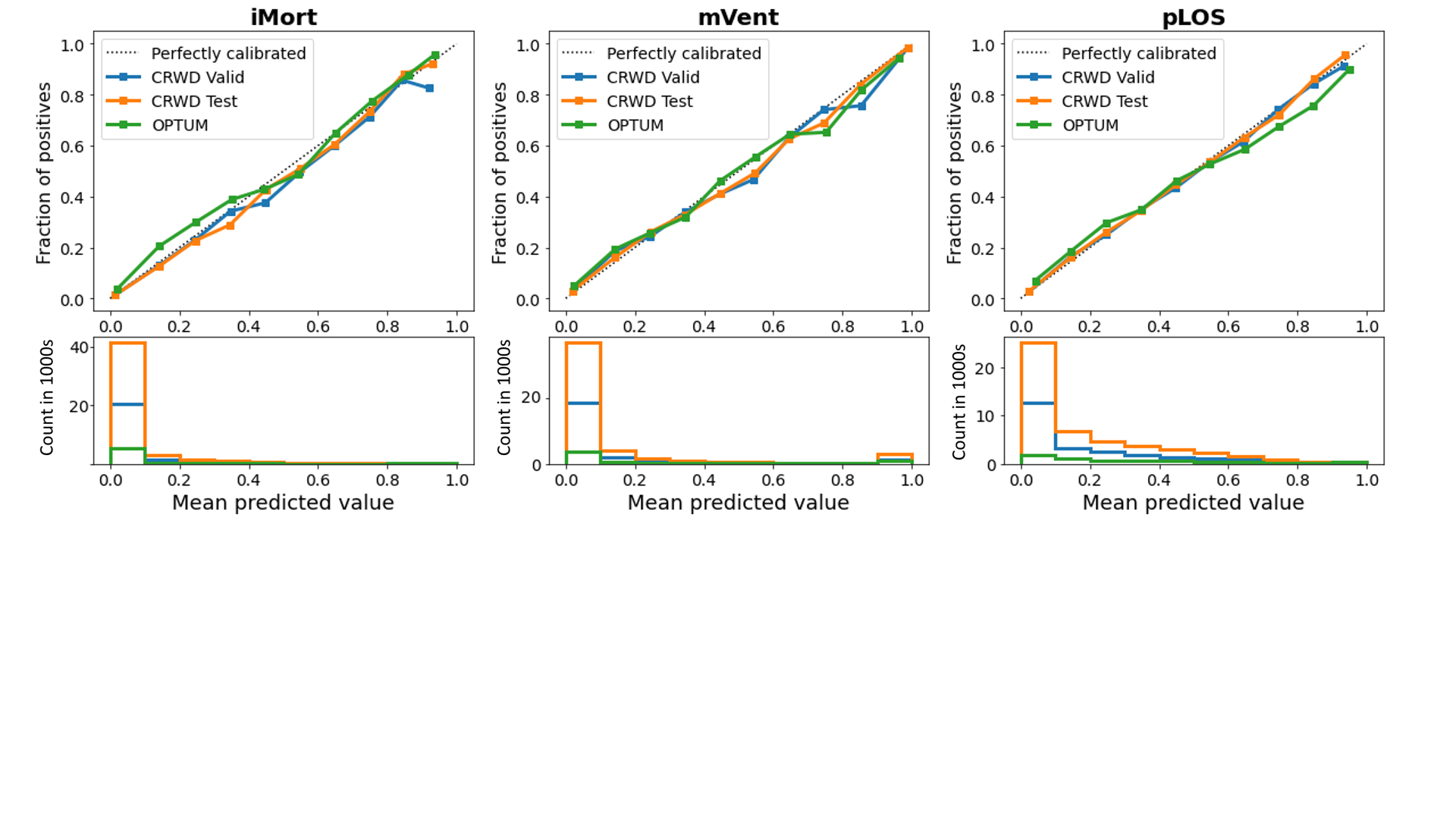

**Supplementary Figure 4. Calibration plots for the CRWD validation set, CRWD multi-hospital test set, and OPTUM test set.**

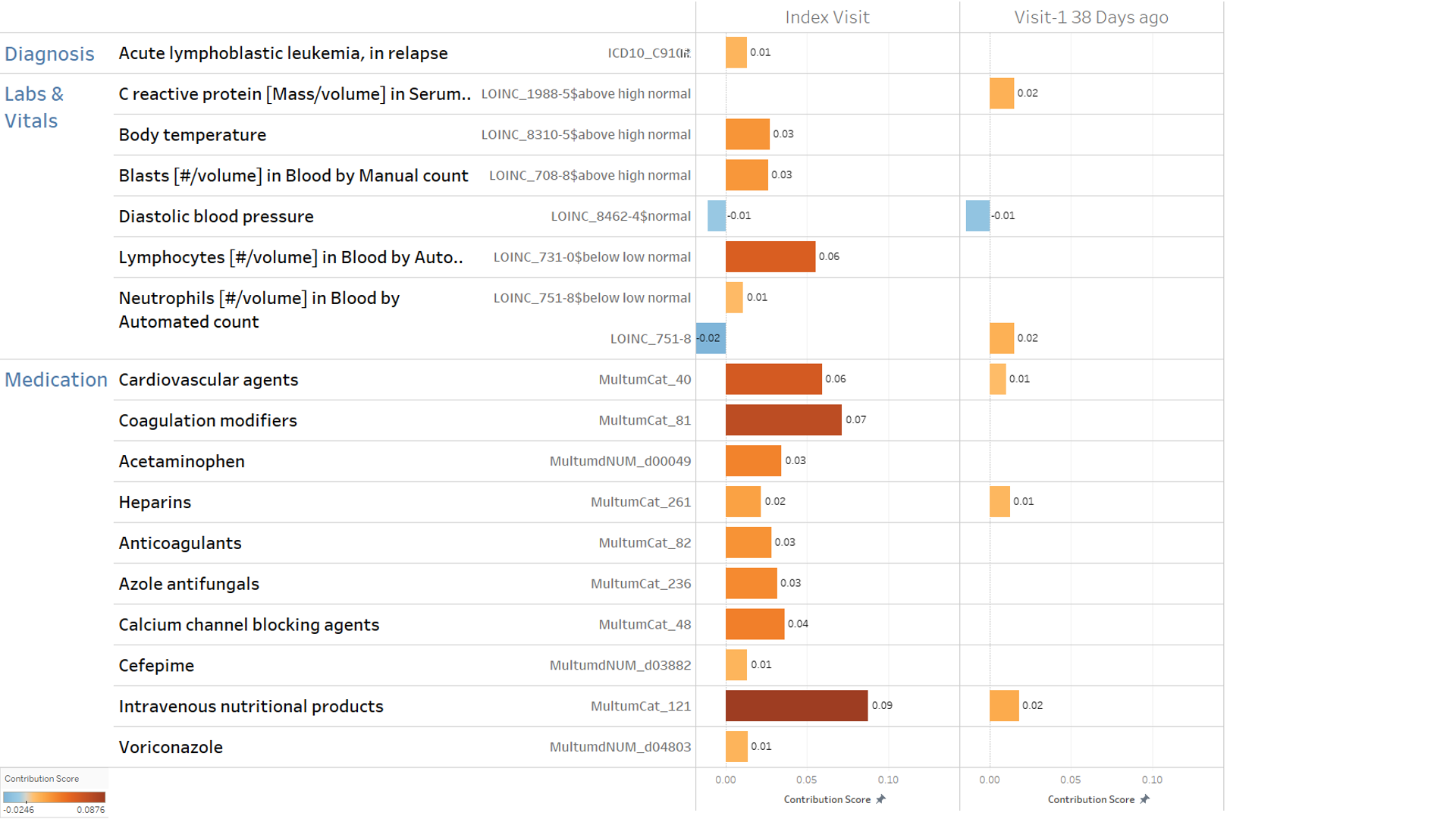

**Supplementary Figure 5. Sample visit level explanation for a true positive pLOS case.**

This is an example patient for whom the CovRNN model correctly predicted, with over 63% probability, would stay more than seven days in the hospital. The bar length and direction represent the contribution score calculated by the integrated gradient that predicts the prolonged hospital stay; i.e., a positive number means a stronger contribution to predicting the prolonged stay. For example, acute lymphoblastic leukemia, in relapse (ICD10_C9102), Blasts in the blood (LOINC_8867-4$above high normal), low lymphocyte counts (LOINC_731-0$below low normal), intravenous nutritional products (MultumCat_121), and coagulation modifiers (MulttumCat_81) are positively correlated to the positive prediction of the prolonged hospital stay, specifically for this patient. Notably, the contribution score can vary at the patient visit level; for example, the neutrophils count (LOINC_751-8) contribution score at an earlier visit was 0.02, whereas at the index visit, it is -0.02, and the patient reported below normal neutrophils and lymphocytes counts, which had a greater contribution to the patient predicted score.
